# Antibiotic-associated neutropenia is marked by depletion of intestinal *Lachnospiraceae* in pediatric patients

**DOI:** 10.1101/2024.04.25.24306386

**Authors:** Josaura Fernandez-Sanchez, Rachel Rodgers, Arushana A. Maknojia, Nusrat Shaikh, Hannah Yan, Marlyd E. Mejia, Hope Hendricks, Robert R. Jenq, Pavan Reddy, Ritu Banerjee, Jeremy M. Schraw, Megan T. Baldridge, Katherine Y. King

**Affiliations:** Department of Pediatrics, Division of Hematology-Oncology, Baylor College of Medicine, Houston, Texas, USA; Texas Children’s Cancer and Hematology Centers, Houston, Texas, USA; Division of Infectious Diseases, Department of Medicine, Washington University School of Medicine, St. Louis, Missouri, USA; Immunology and Microbiology Program, Graduate School of Biomedical Sciences, Baylor College of Medicine, Houston, Texas, USA; Department of Pediatrics, Center for Research Advancement, Baylor College of Medicine, Houston, Texas, USA; Department of Pediatrics, Division of Infectious Diseases, Duke University School of Medicine, Durham, North Carolina, USA; Departments of Genomic Medicine and Stem Cell Transplantation Cellular Therapy, MD Anderson Cancer Center, Houston, Texas; Dan L. Duncan Comprehensive Cancer Center, Baylor College of Medicine, Houston, Texas, USA; Department of Pediatrics, Division of Pediatric Infectious Diseases, Vanderbilt University Medical Center, Vanderbilt University, Nashville, Tennessee, USA; Center for Epidemiology and Population Health, Department of Pediatrics, Baylor College of Medicine, Houston, Texas, USA; Department of Pediatrics, Division of Infectious Diseases, Center for Cell and Gene Therapy, Baylor College of Medicine and Texas Children’s Hospital, Houston, Texas, USA

## Abstract

Hematologic side effects are associated with prolonged antibiotic exposure in up to 34% of patients. Neutropenia, reported in 10-15% of patients, increases the risk of sepsis and death. Murine studies have established a link between the intestinal microbiota and normal hematopoiesis. We sought to identify predisposing factors, presence of microbiota-derived metabolites, and changes in intestinal microbiota composition in otherwise healthy pediatric patients who developed neutropenia after prolonged courses of antibiotics. In this multi-center study, patients with infections requiring anticipated antibiotic treatment of two or more weeks were enrolled. Stool samples were obtained at the start and completion of antibiotics and at the time of neutropenia. We identified 10 patients who developed neutropenia on antibiotics and 29 controls matched for age, sex, race, and ethnicity. Clinical data demonstrated no association between neutropenia and type of infection or type of antibiotic used; however intensive care unit admission and length of therapy were associated with neutropenia. Reduced intestinal microbiome richness and decreased abundance of *Lachnospiraceae* family members correlated with neutropenia. Untargeted stool metabolomic profiling revealed several metabolites that were depleted exclusively in patients with neutropenia, including members of the urea cycle pathway, pyrimidine metabolism and fatty acid metabolism that are known to be produced by *Lachnospiraceae*. Our study confirms a relationship between intestinal microbiota disruption and abnormal hematopoiesis and identifies taxa and metabolites likely to contribute to microbiota-sustained hematopoiesis. As the microbiome is a key determinant of stem cell transplant and immunotherapy outcomes, these findings are likely to be of broad significance.

**Key Points:**

1. Neutropenia occurred in 17% of patients receiving prolonged antibiotic therapy.
2. We found no association between neutropenia and type of infection or class of antibiotic used.
3. Development of neutropenia after prolonged antibiotic treatment was associated with decreased prevalence of *Lachnospiraceae* and Lachnospiraceae metabolites such as citrulline.

## Introduction

More than 236 million antibiotic courses are prescribed every year in the US^1^, many of which represent life-saving therapy for serious infections in children and adults^2^. Yet, antibiotic use can cause significant adverse effects especially when prescribed for prolonged periods. Bone marrow suppression, which can present as anemia, leukopenia with neutropenia and/or thrombocytopenia, occurs in up to 34% of patients receiving antibiotics for two weeks or more ^3–10^. While bone marrow suppression usually improves with early discontinuation of antibiotic therapy^3,8^, shortening antibiotic treatment in response to myelosuppression hampers optimal treatment of the underlying infection. Monitoring for manifestations of bone marrow suppression represents a significant burden of morbidity and healthcare cost. Better understanding the causes of antibiotic-associated bone marrow suppression is critical to developing prevention and treatment strategies that optimize antibiotic use.

Although antibiotic-associated neutropenia has most commonly been reported with beta lactam antibiotics^11^, it has been described as an adverse effect of virtually all classes of antibiotics^4,8,9,12^. The mechanisms underlying antibiotic-associated neutropenia are not well understood, hampering treatment or prevention of this condition. In recent preclinical work, we and others have discovered that antibiotic disruption of the intestinal microbiota plays a critical role in antibiotic-associated cytopenias^13^. In line with those findings, recent studies using murine models demonstrated that prolonged antibiotics can suppress hematopoiesis by affecting the intestinal microbiota^13,15,17–19^. Consistent with this, germ-free (GF) mice have abnormal blood counts and hematopoiesis. Further, recolonization of GF mice with intestinal microbiota results in expansion of the bone marrow myeloid cell pool, an effect that was recapitulated by transferring sterile heat-treated serum or microbial products nucleotide-binding oligomerization domain-containing protein 1 ligand (NOD1L) such as from colonized mice to GF mice^14,15^. Fecal microbiota transplantation (FMT) from young mice to aged mice has also been shown to increase lymphoid differentiation and rejuvenate aged HSCs^16^. Changes in the intestinal microbiota and the metabolites derived from FMT, including *Lachnospiraceae* and tryptophan-associated metabolites, promoted the recovery of hematopoiesis.

Mechanistically, we found that depletion of the intestinal microbiota suppresses type I interferon (IFN) signaling in the bone marrow, whereas oral administration of the microbial metabolites NOD1L or desaminotyrosine (DAT), both of which promote type I IFN signaling, were sufficient to restore normal hematopoiesis and resolve low blood counts in antibiotic-treated mice^20^. These findings support a paradigm in which intestinal microbiota products such as NOD1L and DAT enter the bloodstream and travel to the bone marrow, where they promote the production of basal type I IFN and STAT1 signaling that supports normal hematopoiesis^13^. Altogether, murine studies indicate microbial products in the serum may be necessary to maintain the myeloid cell pool. In support of this paradigm, recent human studies indicate that the state of the intestinal microbiota correlates not only with hematopoietic stem cell transplant (HSCT) outcomes but also with efficacy of cancer immunotherapies^28,30,31^.

While murine studies have elucidated the mechanisms by which the microbiota contribute to normal hematopoiesis, there is a dearth of clinical research studies that validate findings from murine models. Here, we sought to analyze the microbiota and microbiota-derived metabolites in stool samples from pediatric patients receiving prolonged antibiotic therapy. Our study reveals key microbes and microbial metabolites depleted in individuals with antibiotic-associated neutropenia. These findings provide critical mechanistic data that may contribute not only to safe administration of prolonged courses of antibiotics, but may contribute to improved outcomes from HSCT and cancer immunotherapy.

## Methods

### Subjects and Study Design

We carried out a multi-center, non-treatment, non-interventional case-control study approved by Baylor College of Medicine and Affiliated Hospitals’ Institutional Review Board (IRB) and by Vanderbilt University Medical Center’s IRB. Study sites included Texas Children’s Hospital in Houston, Texas and Monroe Carell Jr. Children’s Hospital at Vanderbilt in Nashville, Tennessee. After informed consent was obtained, pediatric patients aged 0 to 18 years were enrolled. Inclusion criteria was admission to the hospital with infections requiring a planned duration of at least two weeks of antibiotic therapy administered either intravenously (IV) or orally (PO). Subjects with a prior history of bone marrow failure or neutropenia, gastrointestinal infections or syndromes, immunodeficiency, malignancy, malnutrition, genetic syndrome known to involve the gastrointestinal or hematopoietic system, antibiotic use during the week preceding hospital admission, or prophylactic antibiotics use were excluded. Complete blood counts were obtained at the discretion of treating providers and monitored by study personnel to identify neutropenia defined as absolute neutrophil count (ANC) of ≤1500/mm^3^. When defining neutropenia, investigators considered a higher threshold of ≤1500/mm^3^ in contrast with that of severe neutropenia (ANC ≤500/mm^3^) perceived as more clinically relevant (1) to allow for earlier identification of cases; and (2) anticipating a rather infrequent CBC monitoring after hospital discharge that potentially prevented identification of severe neutropenia developing in between infrequent blood count checks. Eighty-one patients were enrolled (**Figure 1**).

**Figure 1.**
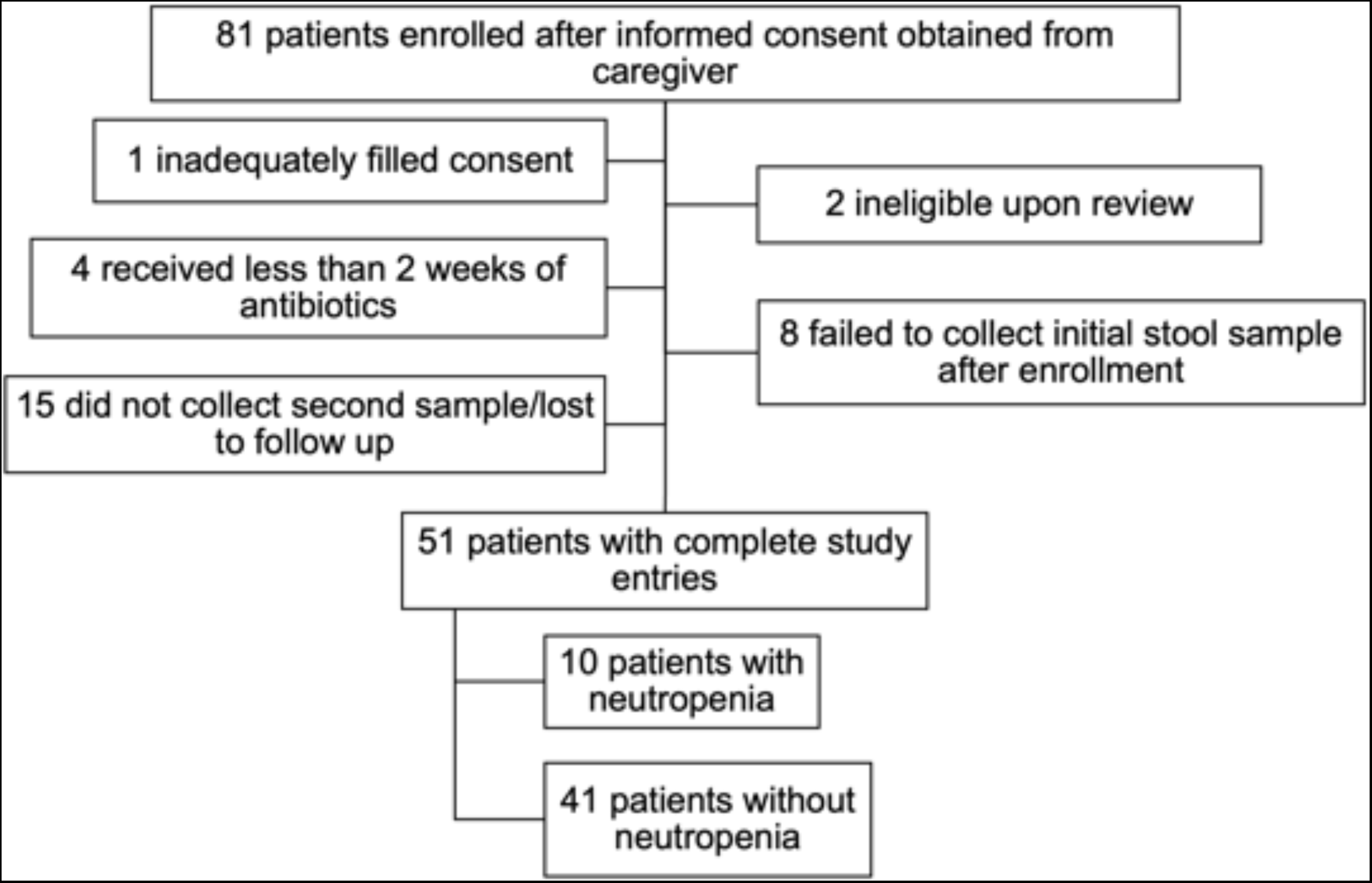
Enrollment Flowchart.

The study included two arms: a *prospective arm*, in which patients were enrolled shortly after admission to the hospital and initiation of antibiotic therapy, and a *retrospective arm*, in which patients who had previously initiated antibiotic therapy were enrolled at time of neutropenia development. Controls were only enrolled in the prospective arm. **Figure 2** illustrates timing of stool samples collection with respect to hospital admission, as well as antibiotic therapy initiation and completion for each subject. Study protocol for the prospective arm was designed so that initial (baseline) stool sample was obtained within 7 days of antibiotic initiation and subsequent (follow-up) stool sample was collected up to 1 week after antibiotic completion or discontinuation due to neutropenia. For the retrospective arm, stool samples were collected at the time of neutropenia development and at least 2-4 weeks after completion of antibiotic therapy and resolution of neutropenia (accounting for “baseline” stool sample). Specific “out of window” stool sample collection timepoints were accepted per investigators discretion for several subjects.

**Figure 2.**
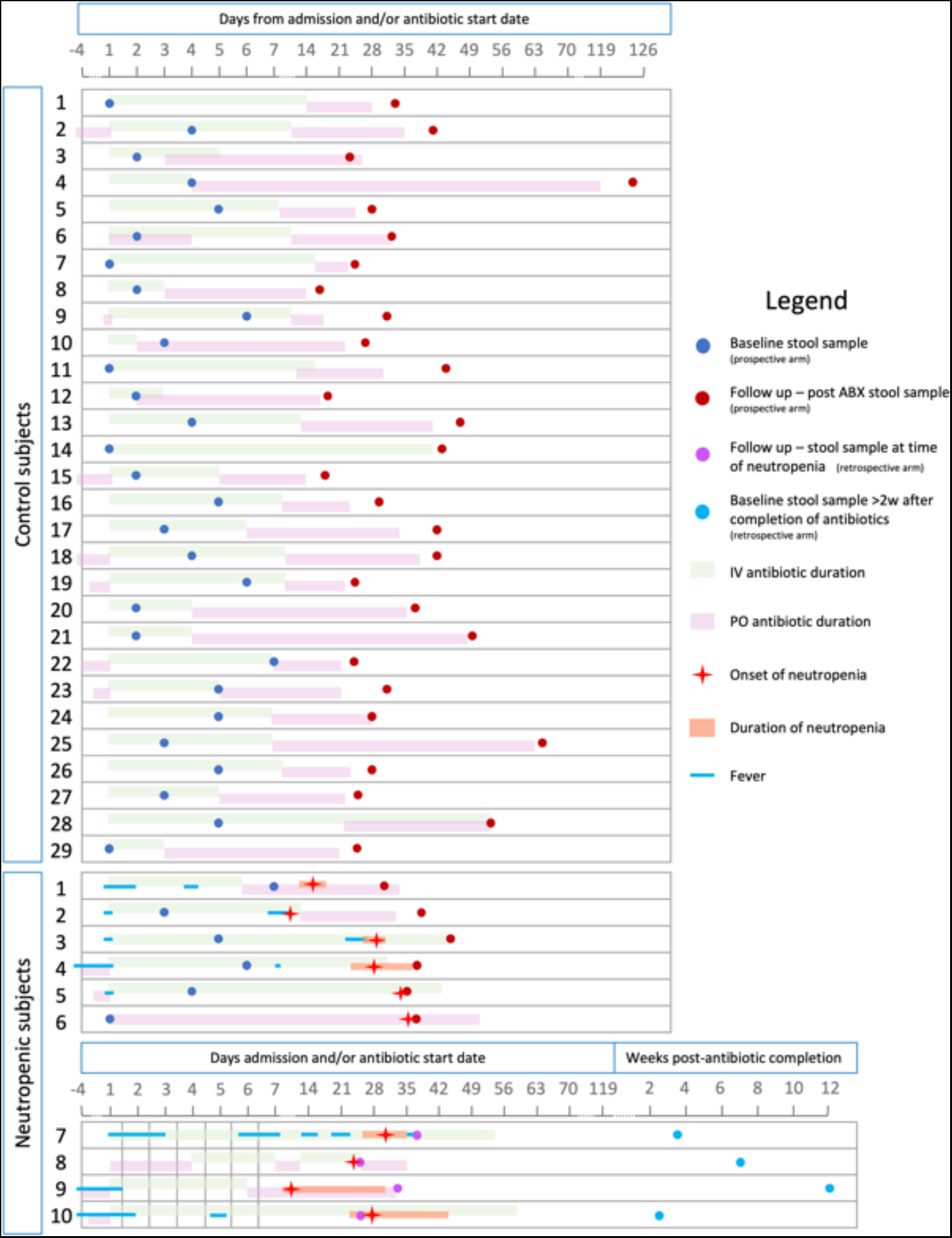
Schematic overview of sampling time and antibiotic treatment duration for each subject. Each study subject is shown on a separate line denoting: intravenous antibiotic treatment (green bar), oral antibiotic treatment (pink bar), baseline stool sample collection (dark and light blue dots for prospective and retrospective arms respectively), stool sample collection after antibiotic treatment (red and purple dots for prospective and retrospective arm, respectively), days of fever (blue line), neutropenia onset (red star) and duration (orange bar). Timelines indicate time since hospital admission in days.

Demographic and clinical information including but not limited to diagnosis, LOS, and laboratory results were collected via electronic medical record review.

We identified ten subjects with neutropenia, including two whose ANC was near 1500 and trending down while antibiotics were continued without reassessment of the CBC, i.e. extrapolation that ANC would have been ≤1500/mm^3^ if blood counts had been re-checked. We used frequency matching on age and race/ethnicity to identify twenty-nine controls. Considering the ethnicity distribution in Houston and Nashville, we combined race and ethnicity to make a single variable (Hispanic, Non-Hispanic White, Non-Hispanic Black, Asian, Native-American).

### Statistical analysis of clinical data

For continuous variables we first identified outliers using the ROUT method at Q=5%; we then performed a Shapiro Wilk normality test with a 0.05 significance. We conducted descriptive analysis by obtaining quartiles and mean/SD/SEM. We used a parametric t-test for normal data (total length of antibiotics, length of PO antibiotics and length of admission) and the Mann-Whitney as non-parametric test for association between continuous and categoric variables. We used chi square testing for most categorical variables, except for number of antibiotic classes used (<2 or >3) where we used Fisher’s exact test.

### Sample collection

Stool samples from participants were collected in sterile plastic containers. The first sample was collected shortly after admission for the prospective arm or at the time of neutropenia for the retrospective arm; the sample was then placed on ice and brought to the laboratory. The second sample was collected at home after discharge, kept in home refrigerators and shipped on ice to the laboratory within hours. Stool samples were processed by investigators upon arrival at the laboratory by aliquoting in 1.5 ml sterile Eppendorf tubes for storage at −80 °C until use. Samples were shipped in dry ice to a central laboratory that performed 16s rRNA gene sequencing and untargeted metabolomics profiling.

### 16S rRNA gene sequencing and untargeted metabolomic analysis

Paired stool samples from individual study subjects were compared by 16S rRNA sequencing and untargeted metabolomic analysis. The supplemental methods provide details about data collection and approaches used to analyze stool samples.

### Data sharing statement

Original 16S rRNA gene sequencing data and untargeted metabolomics data are available through NCBI ENA accession PRJEB72348 and Mendeley datasets https://data.mendeley.com/datasets/38z7h96km4/1, respectively.

## Results

### Patient characteristics

Fifty-one subjects completed study enrollment and sample collection (**Figure 1**). Based on the definition of ANC ≤1500/mm^3^, ten subjects developed neutropenia, four of whom were enrolled in the retrospective arm. Neutropenia occurred in 17% (n=6/47) of subjects in the prospective arm, consistent with previously reported rates in patients on antibiotic therapy for greater than two weeks^3,5,8^. Neutropenia occurred in both male and female subjects and in non-Hispanic White, non-Hispanic Black, Hispanic, and Asian subjects at frequencies that roughly mirrored the population served by the enrolling sites. Twenty-nine subjects without neutropenia were selected as controls by frequency matching for age (within +/- 1 year), sex, and race/ethnicity. The median age was 8.5 years for subjects with neutropenia and 8 years for control subjects, respectively. Baseline demographic characteristics of the selected cohort are shown in **Table 1**.

**Table 1.** Baseline demographic characteristics of subjects treated with long-term antibiotics with and without neutropenia.

|  | Neutropenic | Non-neutropenic | *p* value |
| --- | --- | --- | --- |
| N (total) | 10 | 29 |  |
| Age, median (range) | 8.5 y (6m – 17y) | 8y (6m – 15y) | 0.54 |
| Gender<br>Female, N (%) | 7 (70%) | 14 (48%) | 0.23 |
| Race/Ethnicity<br>Hispanic (%)<br>Non-Hispanic Black (%)<br>Non-Hispanic White (%)<br>Asian (%) | 2 (20%)<br>2 (20%)<br>3 (30%)<br>3 (30%) | 8 (28%)<br>6 (21%)<br>12 (41%)<br>3 (10%) | 0.77 |
| Lowest ANC, median (range) | 895 (460-1570) | 3650 (1666 – 10360) | <0.0001 |

Statistical significance defined as *p* value <0.05 determined by parametric t-test or non-parametric Mann-Whitney test for continuous variables, and Chi square or Fisher's exact test for categorical variables (n= 10 in neutropenic group, 29 in non-neutropenic group). ANC: Absolute neutrophil count.

Other than neonates, all ages were represented in both groups. Infants comprised 20% of subjects with neutropenia and 17% of controls, while children between 1 and 11 years old were slightly more represented in controls compared to the neutropenic group (62% vs. 40%), and adolescents were more represented in the neutropenic group (40% vs 17%). However, no statistically significant correlation was noted between age and neutropenia development.

### Timing of stool sample collection with respect to antibiotic treatment (Figure 2)

A stool sample intended to represent the “baseline” intestinal microbiota was collected shortly after hospital admission for the prospective group, and 2-4 weeks after completion of antibiotic therapy and resolution of neutropenia for the retrospective group. However, given the complexities of study enrollment and sample collection for pediatric patients, timeframes of up to +/- 1 week were established for said sample collections. Alteration of the intestinal microbiome occurs rapidly after antibiotic administration, even after a single dose; and recovery of the microbiome after antibiotic completion can take up to several months^21,22^. In our cohort, there was a wide variation of number of days between first dose of antibiotics and baseline stool sample collection with a median of 4.5 days (range 1-10 days) for the neutropenic group and a median of 3 days (range 1-10 day) for the control group; however, no statistically significant difference was found between groups for this variable (data not shown). For the retrospective arm, “baseline” stool sample was collected a median of 5.25 weeks after completion of antibiotics (range 2.5-12 weeks).

### Factors associated with development of antibiotic-associated neutropenia

Relevant characteristics of the clinical course for subjects in each group are shown in **Table 2**. Neutropenia occurrence was associated with greater total length of antibiotic treatment (*p=0.0021*), intravenous (IV) antibiotic treatment (*p=0.0013*), length of hospital admission stay (LOS) (*p=0.0393*), and admission to the ICU (*p=<0.0001*). Details of the clinical course of each of the neutropenic subjects are provided in **Table S1**.

**Table 2.**
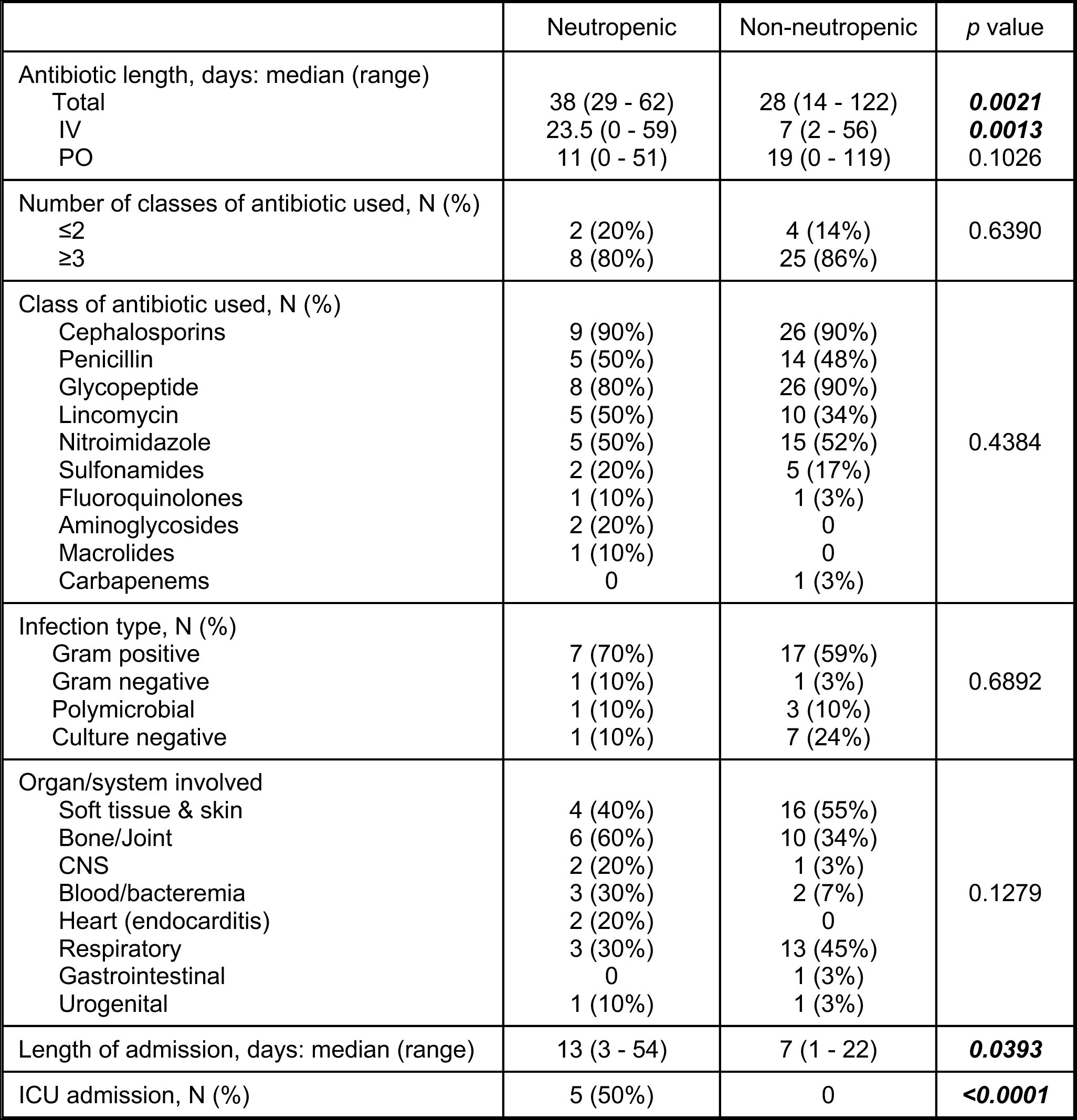
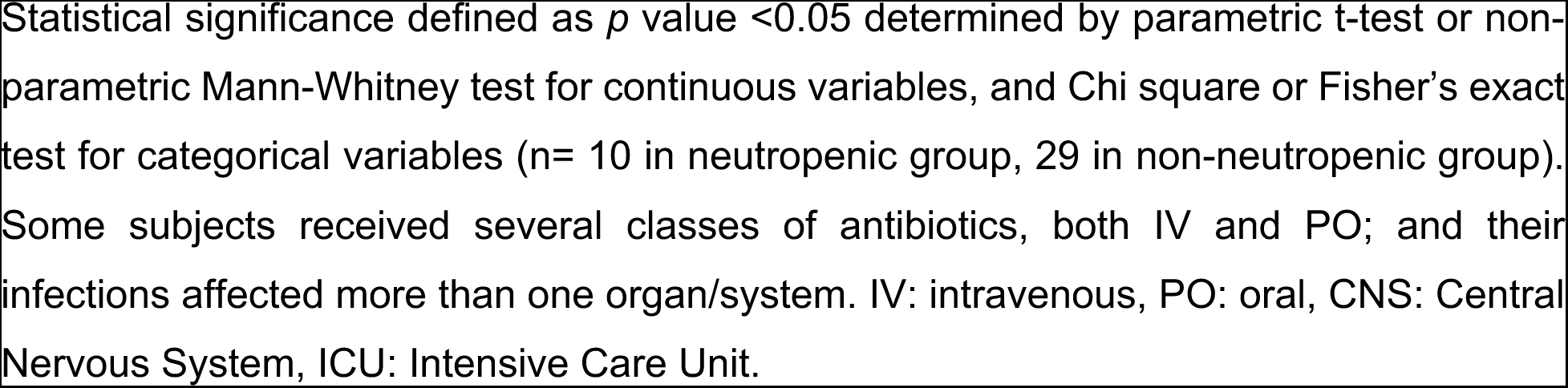
Clinical characteristics of subjects treated with long term antibiotics with and without neutropenia.

|  | Neutropenic | Non-neutropenic | *p* value |
| --- | --- | --- | --- |
| Antibiotic length, days: median (range) |  |  |  |
| Total | 38 (29 - 62) | 28 (14 - 122) | **0.0021** |
| IV | 23.5 (0 - 59) | 7 (2 - 56) | **0.0013** |
| PO | 11 (0 - 51) | 19 (0 - 119) | 0.1026 |
| Number of classes of antibiotic used, N (%) |  |  |  |
| ≤2 | 2 (20%) | 4 (14%) | 0.6390 |
| ≥3 | 8 (80%) | 25 (86%) |  |
| Class of antibiotic used, N (%) |  |  |  |
| Cephalosporins | 9 (90%) | 26 (90%) | 0.4384 |
| Penicillin | 5 (50%) | 14 (48%) |  |
| Glycopeptide | 8 (80%) | 26 (90%) |  |
| Lincomycin | 5 (50%) | 10 (34%) |  |
| Nitroimidazole | 5 (50%) | 15 (52%) |  |
| Sulfonamides | 2 (20%) | 5 (17%) |  |
| Fluoroquinolones | 1 (10%) | 1 (3%) |  |
| Aminoglycosides | 2 (20%) | 0 |  |
| Macrolides | 1 (10%) | 0 |  |
| Carbapenems | 0 | 1 (3%) |  |
| Infection type, N (%) |  |  |  |
| Gram positive | 7 (70%) | 17 (59%) | 0.6892 |
| Gram negative | 1 (10%) | 1 (3%) |  |
| Polymicrobial | 1 (10%) | 3 (10%) |  |
| Culture negative | 1 (10%) | 7 (24%) |  |
| Organ/system involved |  |  |  |
| Soft tissue & skin | 4 (40%) | 16 (55%) | 0.1279 |
| Bone/Joint | 6 (60%) | 10 (34%) |  |
| CNS | 2 (20%) | 1 (3%) |  |
| Blood/bacteremia | 3 (30%) | 2 (7%) |  |
| Heart (endocarditis) | 2 (20%) | 0 |  |
| Respiratory | 3 (30%) | 13 (45%) |  |
| Gastrointestinal | 0 | 1 (3%) |  |
| Urogenital | 1 (10%) | 1 (3%) |  |
| Length of admission, days: median (range) | 13 (3 - 54) | 7 (1 - 22) | **0.0393** |
| ICU admission, N (%) | 5 (50%) | 0 | **<0.0001** |

Statistical significance defined as $p$ value $<0.05$ determined by parametric t-test or non-parametric Mann-Whitney test for continuous variables, and Chi square or Fisher's exact test for categorical variables ( $n= 10$ in neutropenic group, 29 in non-neutropenic group). Some subjects received several classes of antibiotics, both IV and PO; and their infections affected more than one organ/system. IV: intravenous, PO: oral, CNS: Central Nervous System, ICU: Intensive Care Unit.

#### Characteristics of antibiotic regimens

The median duration of antibiotic treatment was 38 days in subjects with neutropenia and 28 days in controls. Analyzing antibiotic administration by mouth (PO) and IV separately indicated that the main difference between groups was in the duration of IV antibiotics **(Table 2)**. Even though the median duration of IV treatment differed significantly between groups, long courses of IV antibiotic treatment up to 50 or more days were noted in both groups, indicating that prolonged IV administration does not always lead to neutropenia. Similarly, subjects that only received PO antibiotics were represented in both groups, indicating that neutropenia can occur after prolonged PO antibiotic therapy alone.

Patients were frequently started on broad spectrum antibiotics upon admission to provide coverage for gram positive, gram negative, and anaerobic pathogens such that the majority of patients in both groups received antibiotics from three or more different classes (80% and 86% in the neutropenia group and controls, respectively). Altogether, subjects received antibiotics from ten different antibiotic classes **(Table 2)**. Cephalosporins, glycopeptides (e.g. vancomycin), nitroimidazoles (e.g. metronidazole), penicillins, and lincomycin (e.g. clindamycin) antibiotics were used often in both groups. Sulfonamides and fluoroquinolones were used less often in both groups, while aminoglycosides, macrolides and carbapenems were used rarely. Whereas clinicians frequently focus on penicillins and sulfonamides (e.g. trimethoprim sulfamethoxazole) as potential neutropenia causes, we found no significant association between the antibiotic class used and neutropenia development in our cohort, likely due to the small size of our cohort and selection bias towards similarly treated infections in otherwise healthy patients.

#### Characteristics of infectious diagnoses

Prolonged antibiotic courses were used to treat a variety of conditions, including skin and soft tissue, bone, joint, and respiratory infections. Osteomyelitis, orbital cellulitis, and sinusitis were the most common diagnoses, with the last two frequently co-occurring. While the numbers were too small to generate any statistically significant association between the organ/system affected and neutropenia development, there were some notable trends. For example, the only two subjects enrolled in our study with infective endocarditis developed neutropenia. In both cases, neutropenia developed after having received antibiotics for more than 30 days, consistent with long treatment durations for endocarditis. Absence of endocarditis among the control cohort is likely related to the overall rarity of infective endocarditis in children^23^. Notably, while there appeared to be an association between bone infections and neutropenia, the inverse was true for lung infections; we speculate this may be explained by treatment duration for bone infections commonly exceeding lung infection treatments by weeks. The central nervous system (mostly affected by intracranial extension of orbital cellulitis), bacteremia (which in all cases co-occurred with other infections), the urogenital system, and the gastrointestinal system were less commonly affected. The involvement of multiple organ systems in the infectious process was not associated with neutropenia in our cohort (**Table 2**).

Individual organisms were isolated in most subjects from both groups; culture negative infection was slightly more common in the control group, although this was not of statistical significance. Overall, infections due to gram positive microorganisms were most common, reflecting the frequency of Staphylococcal and Streptococcal infections in children. Gram positive infections were slightly more common in the neutropenic group than in the control group. We noted no significant association between the isolated microorganism and neutropenia in our cohort.

#### Characteristics of hospital admission

Both LOS and ICU stay were associated with neutropenia **(Table 2)**. Median admission duration was 13 days in the neutropenic group versus 7 days in the control group. Five out of ten subjects with neutropenia required admission to the ICU; however, three of these five subjects were briefly admitted to the ICU for overnight observation after a drainage procedure performed to control their infection and transferred to the acute care floor the next morning after an uneventful ICU course. Detailed review of the ICU admissions showed that the majority of subjects did not have prolonged mechanical ventilation, prolonged NPO (“nothing by mouth”) or enteral feeding periods, prolonged use of renal replacement therapies like hemodialysis or continuous renal replacement therapy, or sedation use. Only one subject required sedation and mechanical ventilation, this persisted for five days after a valvuloplasty procedure. Importantly, no subject developed neutropenia while admitted in the intensive care unit; rather, neutropenia developed days to weeks later. These trends suggest that common factors in the ICU environment did not drive neutropenia development in our cohort. Extended details of the clinical course for each neutropenic subject are available in **Table S1**.

To evaluate for other potential explanations of neutropenia, inflammatory markers, fever and other concomitant signs and symptoms, medications other than antibiotics, neutropenia onset and duration, and overall clinical status at time of neutropenia were assessed. Most subjects became afebrile shortly after admission without fever recurrence; however, two subjects had a brief period of fever preceding neutropenia by ~3 days associated with persistent/worsening infectious abscess identified on imaging that required surgical drainage. All subjects had clinical improvement of infection and overall clinical status at the onset of neutropenia. Consistent with this, inflammatory markers had improved and/or normalized by the time of neutropenia except for one subject for whom this information was not available. No other medications known to cause myelosuppression were administered concurrently with antibiotics. Neutropenia onset occurred at a median of 23 days after antibiotic initiation (range 10-37 days) with neutropenia lasting a median of 8.5 days (range 2-24 days); however, duration was only assessed in 6/10 subjects due to lack of repeat CBCs in the remainder.

### Microbiota diversity contraction and decreased representation of *Lachnospiraceae* taxa are associated with neutropenia

Seventy-six stool samples obtained from 28 control and 10 neutropenic subjects were analyzed by 16S ribosomal RNA gene sequencing. Beta diversity reflecting the total diversity of species represented in each group was assessed by weighted UNIFRAC distance showed significant differences (Adonis *p*=0.031) in the overall composition between groups and timepoints (all samples, **Figure 3A**). However, no significant difference was noted when comparing neutropenic and control subjects at the pre- or post-antibiotic timepoints (not shown), indicating that the major differences in beta diversity were due to antibiotic use, as expected. Changes in community composition by taxa before and after antibiotics in both groups are shown in **Figure 3B**, notably suggesting decreased representation of the Clostridia class (green) after antibiotic treatment only in neutropenic subjects. Alpha diversity assessed by richness reflects the total number of different bacterial taxa (Amplicon Sequence Variants, ASVs) detected within a sample and was significantly decreased in both groups as a result of antibiotic treatment, as expected. The decrease in variability was more significant for subjects with neutropenia (median ASVpre = 124.5, median ASVpost = 29), suggesting increased disruption from baseline compared to the control group (median ASVpre = 111.5, median ASVpost = 50.5); *p= 0.021 vs p= 0.043* respectively) **(Figure 3C)**. Assessment of alpha diversity via Pielou’s Evenness and Shannon Index did not show significance in either group; however, changes in the neutropenic group approached significance.

**Figure 3.**
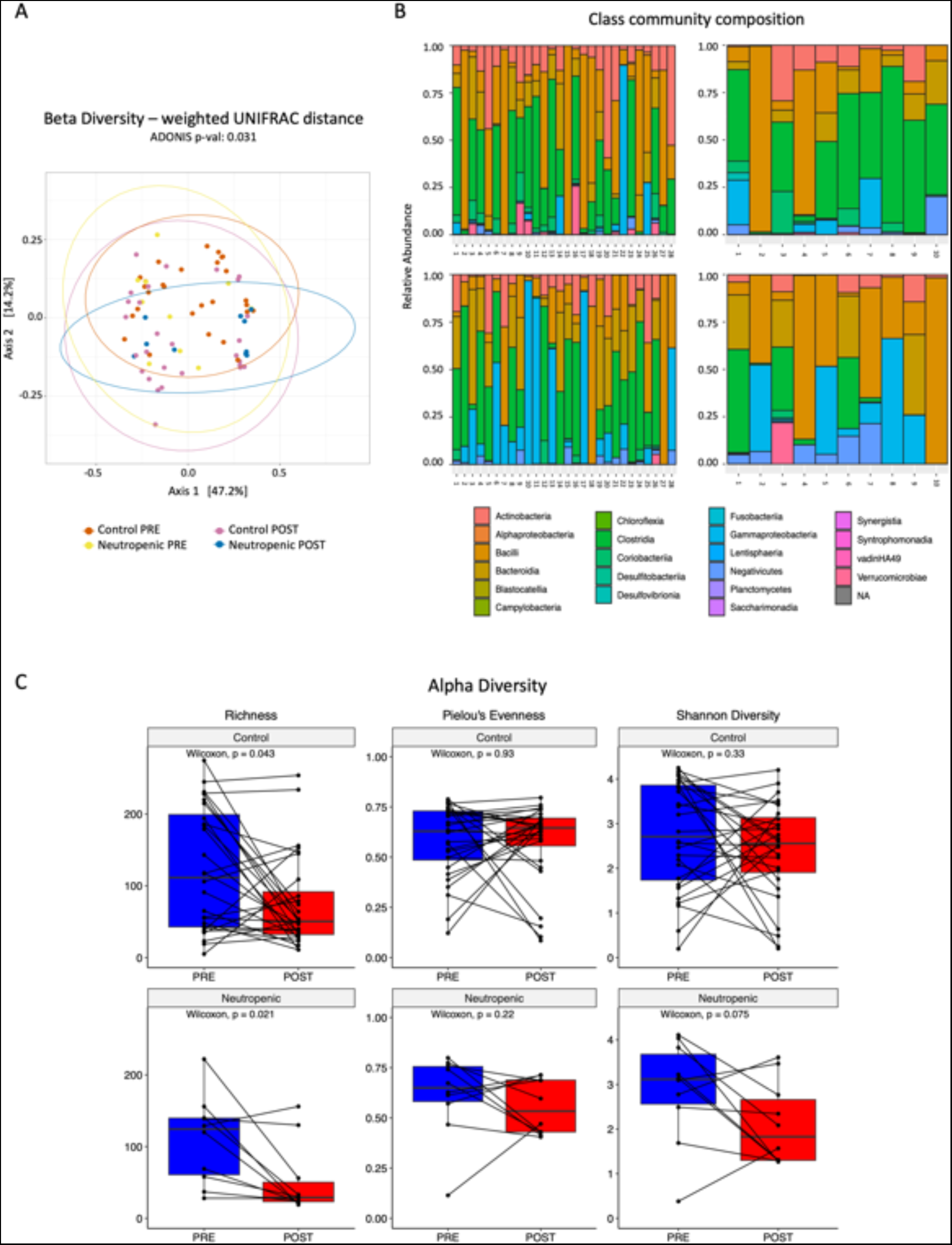
Neutropenic subjects have reduced stool microbiome diversity and decreased predominance of Lachnospiraceae by 16S sequencing. **A.** Beta diversity assessed by weighted UNIFRAC distance. **B.** Class community composition by groups before and after antibiotic treatment. **C.** Alpha diversity assessed by richness, Pielou’s Evenness and Shannon diversity. Statistical significance is defined by *p* <0.05 as determined by Adonis test for beta-diversity, and p <0.05 determined by Wilcoxon test for alpha-diversity analysis (n=10 neutropenic, 28 controls).

Differential analysis of taxa abundance across groups and timepoints was performed with DESeq2 (see supplemental methods). Volcano plots showing differentially abundant taxa between groups and time points are shown in **Figure S1**. **Table 3** shows a list of taxa with significant +log2FC before and after antibiotic treatment uniquely in the neutropenic group (i.e. with decreased representation after antibiotics in neutropenic subjects only), as well as taxa with significant +log2FC after antibiotic treatment in control subjects compared to neutropenic subjects (i.e. with increased representation after antibiotics in control subjects only). Among the 17 taxa included, 10 belong to the Lachnospiraceae family, which includes *Roseburia, Blautia, Anaerostipes, Eubacterium, Ruminococcus*, and *Clostridioides* genera among others. For two of these 17 taxa, namely *Lachnospiraceae [Eubacterium] fissicatena* group (species NA) and *Erysipelotrichaceae [Clostridium] innocuum group* (species NA), the mean abundance significantly decreased after antibiotic treatment by Dunn’s repeat comparisons test (p adj= <0.05) in patients with neutropenia, showing a near complete loss of the latter **(Figure 4A-B)**. There was a trend toward decreased abundance of *Blautia faecis* after antibiotic treatment only in neutropenic subjects (unadjusted p = 0.053; **Figure 4C)**. Additional ground truth plots for taxa that achieved significance only by *unadjusted* p value are shown in **Figures S2A-E**. Comparison of the combined abundances for the ten species belonging to the Lachnospiraceae family shown in **Table 3** demonstrate significantly lower mean abundance in neutropenic subjects after antibiotic treatment but not in controls **(Figure 4D).** Given that relative abundances of bacteria may vary significantly between individuals, we also plotted the data as a change in average abundance between pre and post antibiotic samples for each subject. Plots for *Clostridium innocuum* and *Blautia faecis* are shown in **Figure 4E-F**. Altogether, there was a striking loss of abundance of Lachnospiraceae among neutropenic but not control subjects.

**Figure 4.**
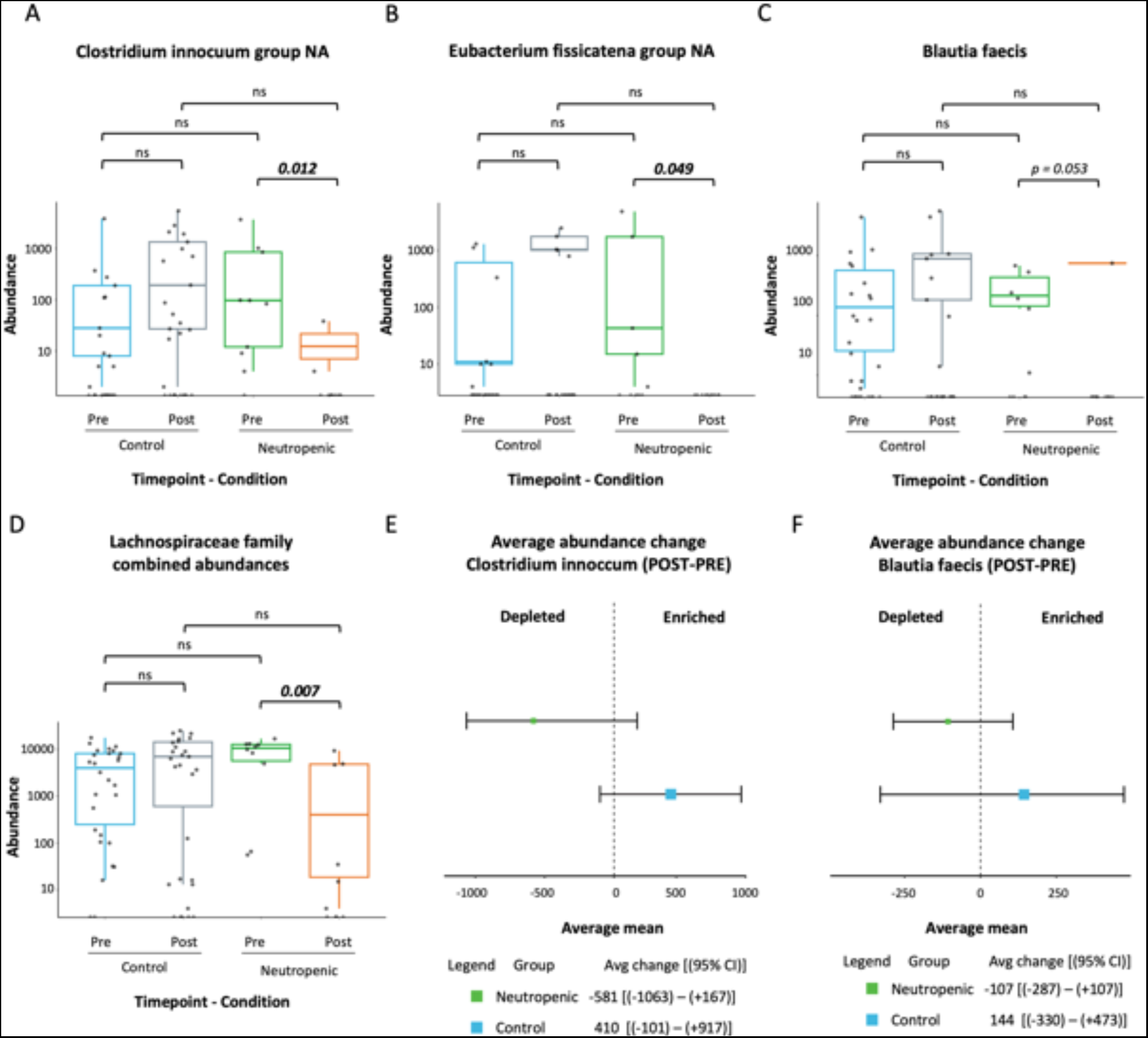
16S rDNA sequencing of stool samples reveals lower mean abundances of several bacterial species after antibiotic treatment in neutropenic subjects. **A-C.** Mean abundances by groups before and after antibiotic treatment. **D.** Combined mean abundances for ten species of the Lachnospiraceae family by groups before and after antibiotic treatment. **E-F.** Forest plots for *Clostridium innocuum* and *Blautia faecis* are shown. Statistical significance is defined by *padj* <0.05 as determined by Dunn’s multiple comparisons test for mean abundances comparisons (n=10 neutropenic, 28 controls).

**Table 3.** Several species among the *Lachnospiraceae* family and others had significant changes in normalized abundance after antibiotic treatment in the neutropenic subjects but not in the controls based on DESeq2 analysis. Table shows species with significant fold changes in abundance after antibiotic treatment in subjects with neutropenia that did not differ or differed in the opposite direction in controls. Two additional taxa of interest were noted to be more abundant after antibiotic treatment in controls compared to neutropenic subjects (bottom).

| Family | Genus | Species | log2FoldChange | p. value | p. adj |
| --- | --- | --- | --- | --- | --- |
| Differentially abundant after antibiotics in neutropenic subjects compared to baseline |  |  |  |  |  |
| Clostridiaceae | Clostridium sensu stricto 1 | NA | 24.5728 | 9.57E-17 | 1.93E-15 |
| Lachnospiraceae | Roseburia | intestinalis | 24.2399 | 2.46E-16 | 3.95E-15 |
| Lachnospiraceae | Roseburia | inulinivorans | 24.2319 | 2.52E-16 | 3.95E-15 |
| Veillonellaceae | Dialister | invisus | 23.0949 | 5.81E-15 | 5.85E-14 |
| Ruminococcaceae | Incertae Sedis | NA | 22.1833 | 6.54E-14 | 5.42E-13 |
| Lachnospiraceae | Coprococcus | comes | 22.0889 | 8.12E-14 | 6.36E-13 |
| Lachnospiraceae | Lachnospiraceae NK4A136 group | NA | 8.3500 | 0.0034 | 0.0221 |
| Peptostreptococcaceae | Romboutsia | NA | 7.4296 | 0.0073 | 0.0445 |
| Lachnospiraceae | Anaerostipes | hadrus | 26.6619 | 3.97E-21 | 5.59E-19 |
| Ruminococcaceae | Ruminococcus | bromii | 25.9072 | 1.92E-18 | 6.76E-17 |
| Lachnospiraceae | [Eubacterium] fissicatena group | NA | 25.5143 | 6.19E-18 | 1.75E-16 |
| Lachnospiraceae | Agathobacter | NA | 23.2200 | 4.10E-15 | 4.60E-14 |
| Enterococcaceae | Enterococcus | NA | 23.2109 | 4.24E-15 | 4.60E-14 |
| Lachnospiraceae | Lachnoclostridium | NA | 22.4440 | 3.30E-14 | 2.91E-13 |
| Erysipelotrichaceae | [Clostridium] innocuum group | NA | 7.1168 | 0.0003 | 0.0024 |
| Differentially abundant after antibiotics in control subjects compared to neutropenic subjects |  |  |  |  |  |
| Lachnospiraceae | Blautia | NA | 21.1322 | 2.49E-10 | 6.95E-09 |
| Lachnospiraceae | Blautia | faecis | 8.5864 | 0.0014 | 0.0358 |

Statistical significance is defined as $p$ . value < 0.05 determined by Wald test (NA = no specie(s) was associated with the sequence by ASV dataset).

### Depletion of specific stool metabolites was associated with increased likelihood of neutropenia development

Untargeted metabolomic profiling was performed on stool samples for all subjects pre- and post-antibiotic therapy. Overall, 1054 metabolites were detected, of which 849 were annotated products of endogenous or bacterial/fungal metabolism. Of these 849 metabolites, 843 were present in both groups before and after antibiotics, indicating high sample quality and adequate sensitivity of detection of all metabolites across groups (**Figure S3A**). Partial Least Squares-Discriminant Analysis (sPLS-DA) demonstrated that metabolomes from subjects with neutropenia and controls generally overlapped both before **(Figure 5A)** and after **(Figure 5B)** antibiotic treatment, although a slight left shift along Component One in baseline samples from neutropenic subjects compared to baseline controls was noted. This raised the intriguing possibility that relative deficiency of some metabolites contributing to Component One may be predictive of the propensity to develop neutropenia upon antibiotic treatment. Metabolites that contributed to Components One and Two are shown in **Figure S3C** and **Table S2**.

**Figure 5.**
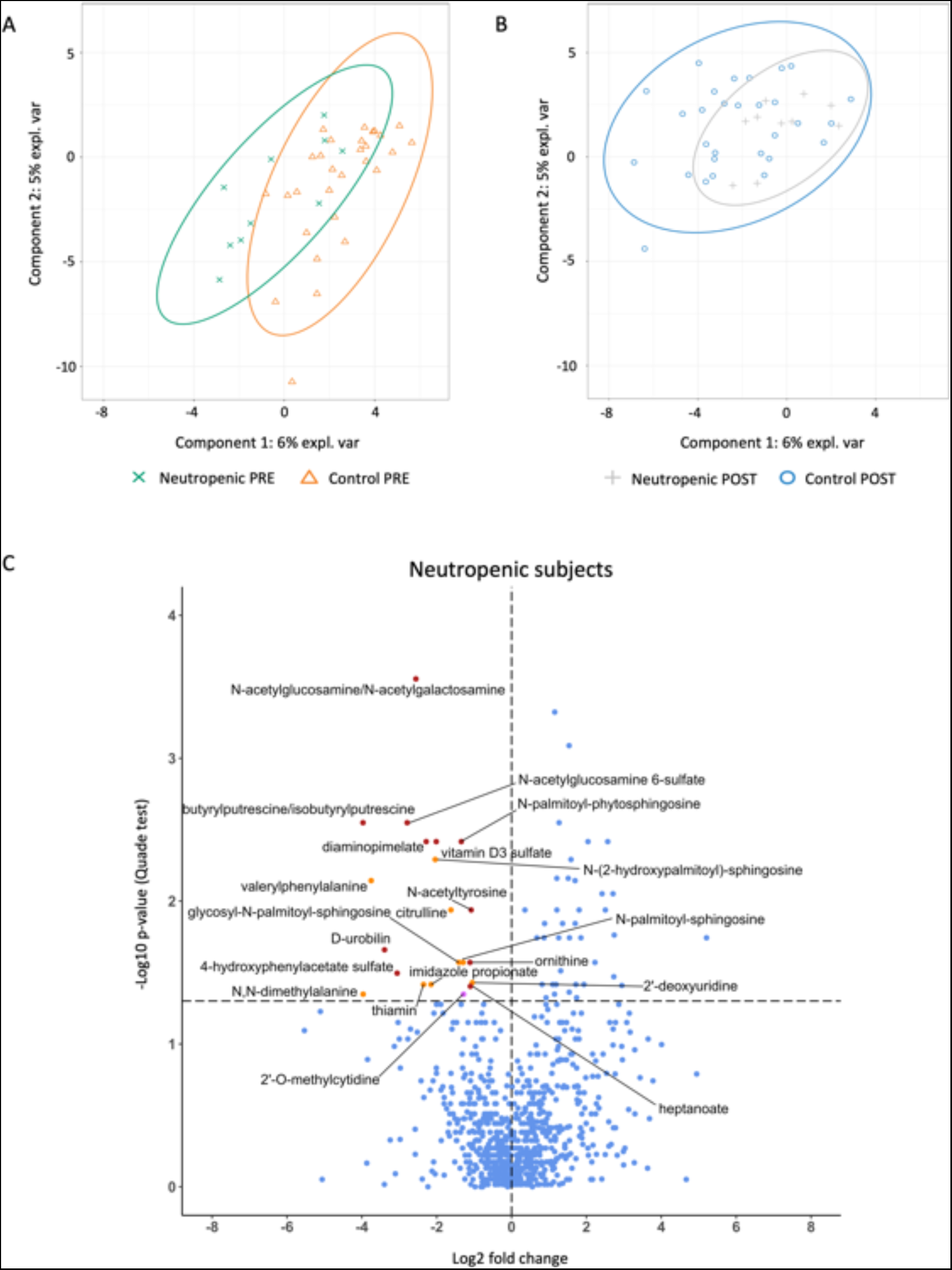
Twenty stool metabolites were depleted after antibiotic treatment in neutropenic subjects but not in controls. Stool samples were assessed by untargeted metabolomic analysis **A-B.** sPLS-DA analysis shows that metabolomes between groups generally overlap between groups before **(A)** and after **(B)** antibiotic treatment. **C.** Volcano plot with annotations shows the twenty-one metabolites that were significantly depleted after antibiotic treatment in neutropenic but not control subjects (orange dots represent metabolites that contribute to sPLS-DA Component One, purple dot for Component Two; remainder of metabolites shown with red dots). Horizontal dotted line indicates p=0.05, statistical significance is defined as p <0.05 determined by Quade test. (n=10 neutropenic, 29 controls)

Using the sPLS-DA component loadings for the metabolites contributing to Components One and Two, logistic regression models were computed to establish odds of neutropenia relative to either metabolite abundance at baseline or the difference in abundance after antibiotic treatment (the latter adjusted for baseline abundance) **(Table 4 and S4)**. According to this analysis, a baseline OR>1 indicates that high baseline abundance (>1 SD) of that metabolite was associated with neutropenia, while a delta OR>1 indicates that an increase in abundance of that metabolite (>1 SD) was associated with neutropenia. At baseline, high abundances of N(6)-methyllysine, alanylproline, phenylacetyltaurine, 3-carboxy-4-methyl-5-pentyl-2-furanpropionate, diacetylspermidine, and N-palmitoyltaurine were each associated with neutropenia. Decreases in N-lactoyl phenylalanine and fructose were associated with lower odds of neutropenia (aOR <1 with p<0.05 indicates >1 SD increased abundance from baseline to follow up sample correlates with lesser odds of neutropenia). Conversely, decreases in 4-ureidobutyrate and citrulline after antibiotic treatment were associated with increased odds of neutropenia (aOR <1 with p<0.05 indicates >1 SD decrease in abundance from baseline to follow up sample correlates with increased odds of neutropenia).

**Table 4.** Several metabolites mapped to PLS-DA components one or two demonstrated significant (p<0.05) associations with neutropenia based on the change in abundance between timepoints adjusted for baseline abundance (aOR change) or based on the baseline abundance (OR baseline).

| Metabolite | Metabolic pathway annotation<sup>1</sup> | PLS-DA component | Component ranking<sup>2</sup> | aOR (95% CI), change<sup>3</sup> | OR (95% CI), baseline<sup>4</sup> |
| --- | --- | --- | --- | --- | --- |
| Citrulline | Urea cycle; Arginine and Proline Metabolism | 2 | 12 | **0.08 (0.01-0.67)** | 1.19 (0.61-2.30) |
| 4-ureidobutyrate | Pyrimidine Metabolism, Uracil containing | 1 | 2 | **0.21 (0.05-0.77)** | 1.59 (0.60-4.19) |
| Fructose | Fructose, Mannose and Galactose Metabolism | 1 | 43 | **2.47 (1.05-5.84)** | 0.93 (0.79-1.09) |
| N-lactoyl phenylalanine | Lactoyl Amino Acid | 1 | 33 | **3.89 (1.07-14.2)** | 0.55 (0.19-1.56) |
| Diacetylspermidine | Polyamine Metabolism | 1 | 32 | 0.76 (0.47-1.24) | **1.46 (1.00-2.14)** |
| Alanylproline | Dipeptide | 1 | 8 | 0.60 (0.24-1.50) | **2.35 (1.07-5.15)** |
| N6-methyllysine | Lysine Metabolism | 1 | 3 | 0.57 (0.22-1.50) | **3.10 (1.16-8.28)** |
| N-palmitoyltaurine | Endocannabinoid | 1 | 34 | 0.47 (0.12-1.91) | **3.40 (1.15-10.1)** |
| 3-carboxy-4-methyl-5-pentyl-2-furanpropionate | Fatty Acid, Dicarboxylate | 1 | 26 | 1.12 (0.67-1.87) | **3.47 (1.1-10.93)** |
| Phenylacetyltaurine | Acetylated Peptides | 1 | 22 | 0.31 (0.03-3.29) | **12.8 (1.13-146.2)** |
| Ribitol | Pentose Metabolism | 1 | 38 | 1.20 (0.69-2.08) | 0.37 (0.12-1.13) |
| N-acetyl-1-methylhistidine | Histidine Metabolism | 1 | 28 | 0.43 (0.16-1.17) | 0.65 (0.18-2.41) |
| Nicotinate | Nicotinate and Nicotinamide Metabolism | 1 | 30 | 1.02 (0.75-1.38) | 1.36 (1.00-1.84) |
| 4-hydroxyphenylacetate | Phenylalanine Metabolism | 1 | 15 | 0.17 (0.03-1.06) | 1.40 (0.78-2.52) |
| N2-acetyllysine | Lysine Metabolism | 2 | 16 | 0.79 (0.49-1.27) | 1.45 (1.00-2.10) |
| N-acetyltaurine | Methionine, Cysteine, SAM and Taurine Metabolism | 1 | 37 | 0.84 (0.61-1.17) | 1.48 (0.96-2.31) |
| 3-hydroxyvalerate | Fatty Acid, Monohydroxy | 2 | 29 | 1.06 (0.52-2.14) | 1.57 (0.92-2.67) |
| Pantothenate | Pantothenate and CoA Metabolism | 1 | 13 | 0.74 (0.39-1.38) | 1.89 (0.98-3.63) |
| Lactosyl-N-stearoyl-sphingosine (d18:1/18:0) | Lactosylceramides | 1 | 47 | 0.60 (0.22-1.59) | 2.54 (0.87-7.42) |
| N6-acetyllysine | Lysine Metabolism | 1 | 7 | 0.84 (0.38-1.87) | 2.66 (0.86-8.21) |
| N6-carboxyethyllysine | Lysine Metabolism | 1 | 6 | 0.83 (0.41-1.68) | 2.99 (0.96-9.33) |
| Tyrosol | Tyrosine Metabolism | 1 | 9 | 0.21 (0.04-1.12) | 3.10 (0.76-12.6) |
| Beta-sitosterol | Sterol | 1 | 45 | 1.09 (0.40-2.98) | 3.44 (0.95-12.5) |
| 2-hydroxysebacate | Fatty Acid, Dicarboxylate | 1 | 18 | 0.73 (0.29-1.81) | 4.51 (0.84-24.2) |

Abbreviations: aOR=adjusted odds ratio; OR=odds ratio; CI=confidence interval PLS-DA = partial least squares-discriminant analysis.
<sup>1</sup>Per Metabolon.
<sup>2</sup>From 1 (highest) to 50 (lowest), based on the absolute value of the loading score.
<sup>3</sup>OR and 95% CI of neutropenia at the post-treatment timepoint per one standard deviation increase in metabolite abundance from the pre-treatment (baseline) timepoint to the post-treatment (follow-up) timepoint. Adjusted for baseline metabolite abundance.
<sup>4</sup>OR and 95% CI of neutropenia at the post-treatment timepoint per one standard deviation increase in metabolite abundance at the pre-treatment (baseline) timepoint

All metabolites associated with neutropenia by baseline or change in abundance in this logistic regression analysis were part of Component One except for citrulline, which is part of Component Two. To further understand relationships between metabolites altered in the study, we performed over-representation analysis (ORA) using sPLS-DA component loadings. This analysis indicated that Component One was enriched for metabolites involved in pantothenate and coenzyme A biosynthesis, whereas Component Two demonstrated enrichment for metabolites involved in sphingolipid metabolism, pyrimidine metabolism (upstream of urea cycle), butyrate metabolism, and thiamine metabolism **(Figure S3)**.

### Several metabolites were uniquely depleted after antibiotic treatment in subjects with neutropenia

We further conducted a Quade test to identify changes in specific stool metabolites before and after antibiotic treatment in neutropenic versus control subjects. This analysis capitalizes on the paired longitudinal sampling in our study by focusing on changes in abundance rather than absolute abundances. Quade analysis revealed that twenty-one metabolites were significantly depleted after antibiotic treatment in subjects with neutropenia but not in controls **(Figure 5C).** These metabolites include the bacterial cell wall component N-acetylglucosamine, the urea cycle intermediates citrulline and ornithine; as well as the pyrimidine metabolism intermediary 2’-deoxyuridine. Results of the statistical analysis by Quade test for these metabolites are shown in **Table S3**. We observed that several metabolites associated with neutropenia were differentially abundant by Quade test as well as in the logistic regression analysis of sPLS-DA Components One and Two. Namely, urea cycle intermediary citrulline was significantly depleted after antibiotic treatment only in neutropenic subjects **(Figure 5C)**, which was in turn associated with increased risk of neutropenia **(Table 4 and S4)**. Meanwhile, pyrimidine and butyrate metabolism surfaced in both the ORA analysis and the logistic regression of PLS-DA analysis, and members of the pyrimidine synthesis pathway have been noted in our prior murine studies **(Figure S3)**^20^. Interestingly, citrulline and butyrate levels have been reported to correlate with prevalence of Lachnospiraceae in prior studies; therefore loss of Lachnospiraceae specifically among neutropenic subjects may predict loss of abundance of these metabolites associated with neutropenia^24^.

## Discussion

In this study, we enrolled subjects receiving prolonged courses of antibiotics and compared clinical and biological characteristics between subjects who developed neutropenia with controls who maintained normal neutrophil counts. There was a 17% neutropenia rate in the prospective arm of our study, validating the incidence previously reported in the literature^3,5,8^. We found that longer exposure to antibiotics correlated with neutropenia development. A wide array of antibiotic classes, infective microorganisms, and organ systems infected were represented in our cohort. While recognizing the power limitation of our study, we did not find a clear association between these characteristics and neutropenia development. Crucially, neutropenia was associated with contraction of gastrointestinal microbiome diversity, and taxonomic analysis revealed that decreased abundance of several species within the *Lachnospiraceae* family was associated with neutropenia after prolonged antibiotic treatment. Several stool metabolites were depleted only in subjects with neutropenia, including bacterial cell wall component N-acetylglucosamine/N-acetylgalactosamine and urea cycle metabolites citrulline and ornithine. Additionally, sPLS-DA and ORA analysis suggest that decreased abundances of 4-ureidobutyrate and citrulline after antibiotics were associated with increased odds of neutropenia. Overall, these clinical data from a relatively small cohort of individuals reinforce the concept that (1) commensal bacteria support steady-state hematopoiesis by supporting the production of specific metabolites and (2) their disruption by prolonged antibiotic exposure may result in antibiotic-associated cytopenias.

The literature has long described bone marrow suppression as a result of prolonged antibiotic therapy^25–27^. This continues to be a clinical problem because bone marrow suppression interferes with treatment of infections and increases the risks of morbidity and mortality, as well as healthcare costs^17^. Patients with oncologic conditions or who require HSC transplantation are often at increased risk of sepsis due to prolonged neutropenia that results from chemotherapeutic agents. Additionally, frequent use of broad-spectrum antibiotics in these populations, both for prophylactic and therapeutic purposes, diminishes microbial diversity, which has been linked to worse transplant outcomes with increased mortality, increased graft-versus-host disease rates, and increased risk of graft failure and infections^28–32^. Pre-clinical findings from murine studies support the idea of microbiota-sustained steady-state hematopoiesis and our recent work establishes that type I IFN-STAT1 signaling is critical in this interaction^20^. Our current study focuses on adding to these pre-clinical data in the clinical setting by analyzing: (1) the intestinal microbiome in stool samples from otherwise healthy human patients; (2) disruption patterns in the microbiome after antibiotic therapy; and microbiome disruption’s correlation to the developing antibiotic-associated neutropenia.

We identified associations between neutropenia and the total length of antibiotic therapy, length of intravenous antibiotic therapy, length of admission to the hospital and admission to the intensive care unit **(Table 2)**. Previous reports concur with the increased risk of neutropenia with longer treatment courses, as we saw in our cohort^33,34^. While standard intravenous and oral dosing for many common antibiotics (e.g. clindamycin) results in similar blood concentrations, prolonged oral antibiotic administration did not correlate with neutropenia in our cohort. One possible explanation is that more severe infections that tend to require a longer IV treatment duration also have a greater impact on hematopoiesis. In particular, we wonder whether osteomyelitis’ impact on bone marrow physiology, when combined with microbiota-mediated effects, may increase cytopenia risks. Even though some studies suggest that PO antibiotics impact the gut microbiome in a direct or local manner, and that IV antibiotics have an impact only through biliary excretion^35^; recent studies typically link both IV and PO antibiotics with intestinal microbiota changes, including the well-studied influence of intrapartum IV antibiotic prophylaxis in neonatal microbiota composition^21,22,36,37^. Our detailed review did not reveal ICU-specific events that could further disrupt the intestinal microbiota and explain the correlation noted in our cohort. Further studies in larger cohorts could help elucidate whether common aspects of admission to the ICU, such as stress, anesthesia, or nutritional status increase the likelihood of antibiotic-associated neutropenia. While alternative mechanisms of neutropenia such as drug-induced, immune mediated, or sepsis and cytokine storm-induced myelosuppression are plausible^11,12^; our comprehensive review of fever curve, concomitant symptoms, inflammatory markers, clinical signs and symptoms, as well as overall improvement of infection (versus lack thereof) suggest that those mechanisms are not at play in our cohort.

We did not identify a correlation between neutropenia and antibiotic class, number of antibiotic classes used, organ/system involved or isolated microorganisms. Although beta lactams are classically recognized to cause neutropenia, several other classes of antibiotics were represented in our neutropenic cohort, which coincides with what has been previously described in the literature ^4,8,9,12^. Even though no specific class of microorganism was associated with neutropenia, the predominance of gram-positive infections reflects the overall high prevalence of these infections in the pediatric population^38,39^. However, our study was not powered to detect such differences and larger prospective studies will be informative to address these questions.

As expected, richness was decreased in both groups after antibiotic treatment. However, this depletion was more significant in subjects with neutropenia, suggesting that more profound microbiome disruption alters steady state hematopoiesis and increases the likelihood of developing cytopenias. Interestingly, differential taxa abundance analysis showed decreased predominance of several species within the *Lachnospiraceae* family and other Clostridia **(Table 3)**. This family is a phylogenetically and morphologically heterogeneous taxon belonging to the clostridial cluster XIVa of the phylum Firmicutes and is among the top short chain fatty acid (SCFA) producers. Within the *Lachnospiraceae* family, *Blautia, Coprococcus, Dorea, Lachnospira, Oribacterium, Roseburia,* and *L-Ruminococcus* are the main genera that metagenomic analyses have detected in the human intestinal microbiota^40^.

Changes in *Lachnospiraceae* abundance have been linked to both health and disease, including metabolic syndrome, obesity, diabetes, liver diseases, inflammatory bowel disease, and chronic kidney disease^40^. Most notably, Jenq *et al* noted that depleting the genus *Blautia* was associated with worse outcomes after HSC transplantation and higher rates of GVHD^29^. Interestingly, in our cohort, *Blautia faecis* was significantly more represented by DESEQ analysis in samples from control subjects after antibiotics when compared to samples from neutropenic subjects after antibiotics **(Table 3)**.

We identified many metabolites that were depleted after antibiotic treatment only in subjects with neutropenia **(Figure 5C, Table S3)** and that correlated with increased likelihood of neutropenia **(Table 4)** based on regression models performed with sPLS-DA component loadings. Notably, several of these metabolites were also detected in serum and/or stool from leukopenic mice in our prior murine work, with significant changes in pyrimidine metabolites such as orotidine and 3-Amino-2-piperidone in the serum of leukopenic mice^20^. The sPLS-DA analysis of metabolomes showed a slight shift in the metabolomic profile of subjects who went on to develop neutropenia **(Figure 5A-B)** suggesting lower abundance of certain Component One metabolites at baseline may predispose to antibiotic-associated neutropenia.

Excitingly, a number of metabolites identified in our metabolomic study are related to bacterial species identified orthogonally through our microbiome analysis. Among these, low plasma levels of citrulline have been associated with decreased abundance of Lachnospiraceae in severely malnourished patients with anorexia nervosa receiving enteral nutrition^41^. Another study linked antibiotic use, lower bacterial abundance, and plasma citrulline levels with worse response to treatment with Nivolumab in a cohort of patients with non-small cell lung cancer^42^. We also observed decreased 4-ureidobutyrate in neutropenic subjects, an intermediary of pyrimidine metabolism. Cytidine supplementation has been reported to improve dyslipidemia in obese mice via modulation of the microbiome with restoration of short chain fatty acid (SCFA) producing bacteria, including *Lachnospiraceae*^40,43^. Butyrate-producing bacteria regulate various gut-associated functions, including energy production, regulating gut barrier integrity by limiting pro-inflammatory cytokine production, and even inhibiting oncogenic pathways^44,45^. These bacteria also help maintain an anaerobic environment and prevent colonization of other pathogenic bacteria. A study by Hayase *et al*. reported reduced fecal levels of butyrate in antibiotic-treat HSC transplant (HSCT) recipient mice that suffered from aggravated GVHD^46^.

Our study was limited by a small sample size of neutropenic subjects. Importantly, the study design allowed for stool samples intended to represent the baseline intestinal microbiota to be collected after several days of antibiotic exposure which affects our ability to interpret our findings at baseline. However, the lack of significant differences in beta diversity analysis between groups, as well as the overlapped detection of 843 out of 849 metabolites in all groups suggests that baseline samples were affected similarly by antibiotic exposure. Additionally, some of the antibiotic classes were rarely used to treat subjects in our cohort - specifically aminoglycosides, macrolides and carbapenems. In the future, larger sample sizes may increase our ability to detect differences between groups. While we analyzed the total length of antibiotic therapy and the number of classes used, a more in-depth analysis accounting for the treatment duration of each antibiotic and timing of administration with respect to neutropenia development could unveil specific associations.

Altogether, current studies support the relevance of the intestinal microbiota in normal hematopoiesis as well as in outcomes from HSCT and immunotherapy outcomes. To our knowledge, this is the first study to analyze microbiome composition changes and intestinal microbiota-derived metabolites after prolonged antibiotic therapy and their relationship to antibiotic-associated neutropenia in a pediatric population. Our study provides the first evidence for the importance of Lachnospiraceae and its metabolites in steady state hematopoiesis and reveals bacterial species and metabolites of interest that may be predictive of neutropenia risk or used therapeutically important in preventing and treating neutropenia. Improved definition and validation of these findings will help us develop risk prediction models that will identify patients at risk of developing antibiotic-associated neutropenia. This will be of particular importance in immunologically fragile populations such as HSCT recipients for whom neutropenia is especially dangerous.

## Data Availability

All data produced in the present study are available upon reasonable request to the authors or online at: https://www.ebi.ac.uk/ena/browser/view/PRJEB72348 and https://data.mendeley.com/datasets/38z7h96km4/1

https://data.mendeley.com/datasets/38z7h96km4/1

## Acknowledgements

The authors thank the Division of Pediatrics Center for Research Advancement at Texas Children’s Hospital - Baylor College of Medicine for their support with subject screening and enrollment and the Center for Metagenomics and Microbiome Research at Baylor College of Medicine. We acknowledge Enrico Moiso PhD for his contribution with the initial analysis of 16S rRNA sequencing data.

This project was supported by the National Institutes of Health (NIH) R01 AI141716 (M.T.B. and K.Y.K.), R35HL155672 (K.Y.K.), F31HL168921 (A.M.), T32GM136554 (A.M.), F31HL147514 A1 (H.Y.) and F31AI167538 (M.E.M). J.F.S. was supported by Baylor College of Medicine Comprehensive Cancer Training Program via Cancer Prevention & Research Institute of Texas Training Award RP210027.

## Authorship and conflicts statement

H.Y., K.Y.K., and M.T.B. designed the study protocol; R.J. and P.R. provided critical insights; J.F.S. and K.Y.K. proposed and reviewed subsequent protocol amendments; H.Y., J.F.S., and N.S. enrolled subjects and, together with A.M., coordinated sample collection and processing; H.H. and R.B. enrolled subjects at participating site; J.F.S. collected, analyzed and interpreted clinical data; J.S. analyzed and interpreted metabolomics results; M.E.M, R.R. and R.J. analyzed and interpreted 16S rDNA sequencing results; J.F.S. and K.Y.K wrote the manuscript; all authors have read and agreed to the published version of the manuscript.

The authors declare no competing financial interests.

## Notes

### Competing Interest Statement

The authors have declared no competing interest.

### Funding Statement

This study was funded by R01AI141716, R35HL155672, F31HL168921, F31HL147514, T32GM136554, and R31AI167538.

### Author Declarations

The IRBs of Baylor College of Medicine and Vanderbilt University gave ethical approval for this work.

## References

1. CDC. Antibiotic Use in the United States, 2022 Update: Progress and Opportunities.

2. Tice AD, Rehm SJ, Dalovisio JR, et al. Practice guidelines for outpatient parenteral antimicrobial therapy. IDSA guidelines. Clin Infect Dis. 2004;38(12):1651–1672. doi:10.1086/420939

3. Gomez M, Maraqa N, Alvarez A, Rathore M. Complications of outpatient parenteral antibiotic therapy in childhood. Pediatr Infect Dis J. 2001;20(5):541–543. doi:10.1097/00006454-200105000-00015

4. Same RG, Hsu AJ, Cosgrove SE, et al. Antibiotic-Associated Adverse Events in Hospitalized Children. J Pediatric Infect Dis Soc. 2021;10(5):622–628. doi:10.1093/jpids/piaa173

5. Olson SC, Smith S, Weissman SJ, Kronman MP. Adverse Events in Pediatric Patients Receiving Long-Term Outpatient Antimicrobials. J Pediatric Infect Dis Soc. 2015;4(2):119–125. doi:10.1093/jpids/piu037

6. Furtek KJ, Kubiak DW, Barra M, Varughese CA, Ashbaugh CD, Koo S. High incidence of neutropenia in patients with prolonged ceftaroline exposure. J Antimicrob Chemother. 2016;71(7):2010–2013. doi:10.1093/jac/dkw062

7. LaVie KW, Anderson SW, O’Neal HR, Rice TW, Saavedra TC, O’Neal CS. Neutropenia Associated with Long-Term Ceftaroline Use. Antimicrob Agents Chemother. 2016;60(1):264–269. doi:10.1128/AAC.01471-15

8. Madigan T, Banerjee R. Characteristics and outcomes of outpatient parenteral antimicrobial therapy at an academic children’s hospital. Pediatr Infect Dis J. 2013;32(4):346–349. doi:10.1097/INF.0b013e31827ee1c2

9. Tamma PD, Avdic E, Li DX, Dzintars K, Cosgrove SE. Association of Adverse Events With Antibiotic Use in Hospitalized Patients. JAMA Intern Med. 2017;177(9):1308–1315. doi:10.1001/jamainternmed.2017.1938

10. Fernandes P, Milliren C, Mahoney-West HM, Schwartz L, Lachenauer CS, Nakamura MM. Safety of Outpatient Parenteral Antimicrobial Therapy in Children. Pediatr Infect Dis J. 2018;37(2):157–163. doi:10.1097/INF.0000000000001716

11. Cimino C, Allos BM, Phillips EJ. A Review of β-Lactam-Associated Neutropenia and Implications for Cross-reactivity. Ann Pharmacother. 2021;55(8):1037–1049. doi:10.1177/1060028020975646

12. Lam PW, Leis JA, Daneman N. Antibiotic-Induced Neutropenia in Patients Receiving Outpatient Parenteral Antibiotic Therapy: a Retrospective Cohort Study. Antimicrob Agents Chemother. 2023;67(3):e0159622. doi:10.1128/aac.01596-22

13. Yan H, Baldridge MT, King KY. Hematopoiesis and the bacterial microbiome. Blood. 2018;132(6):559–564. doi:10.1182/blood-2018-02-832519

14. Balmer ML, Schürch CM, Saito Y, et al. Microbiota-derived compounds drive steady-state granulopoiesis via MyD88/TICAM signaling. J Immunol. 2014;193(10):5273–5283. doi:10.4049/jimmunol.1400762

15. Iwamura C, Bouladoux N, Belkaid Y, Sher A, Jankovic D. Sensing of the microbiota by NOD1 in mesenchymal stromal cells regulates murine hematopoiesis. Blood. 2017;129(2):171–176. doi:10.1182/blood-2016-06-723742

16. Zeng X, Li X, Li X, et al. Fecal microbiota transplantation from young mice rejuvenates aged hematopoietic stem cells by suppressing inflammation. Blood. 2023;141(14):1691–1707. doi:10.1182/blood.2022017514

17. Josefsdottir KS, Baldridge MT, Kadmon CS, King KY. Antibiotics impair murine hematopoiesis by depleting the intestinal microbiota. Blood. 2017;129(6):729–739. doi:10.1182/blood-2016-03-708594

18. Han H, Yan H, King KY. Broad-Spectrum Antibiotics Deplete Bone Marrow Regulatory T Cells. Cells. 2021;10(2). doi:10.3390/cells10020277

19. Kennedy EA, King KY, Baldridge MT. Mouse Microbiota Models: Comparing Germ-Free Mice and Antibiotics Treatment as Tools for Modifying Gut Bacteria. Front Physiol. 2018;9:1534. doi:10.3389/fphys.2018.01534

20. Yan H, Walker FC, Ali A, et al. The bacterial microbiota regulates normal hematopoiesis via metabolite-induced type 1 interferon signaling. Blood Adv. 2022;6(6):1754–1765. doi:10.1182/bloodadvances.2021006816

21. Doan T, Liu Z, Sié A, et al. Gut Microbiome Diversity and Antimicrobial Resistance After a Single Dose of Oral Azithromycin in Children: A Randomized Placebo-Controlled Trial. Am J Trop Med Hyg. 2024;110(2):291–294. doi:10.4269/ajtmh.23-0651

22. Gough EK. The impact of mass drug administration of antibiotics on the gut microbiota of target populations. Infect Dis Poverty. 2022;11(1):76. doi:10.1186/s40249-022-00999-5

23. Vicent L, Luna R, Martínez-Sellés M. Pediatric Infective Endocarditis: A Literature Review. J Clin Med. 2022;11(11). doi:10.3390/jcm11113217

24. Vacca M, Celano G, Calabrese FM, Portincasa P, Gobbetti M, De Angelis M. The Controversial Role of Human Gut Lachnospiraceae. Microorganisms. 2020;8(4). doi:10.3390/microorganisms8040573

25. Olaison L, Belin L, Hogevik H, Alestig K. Incidence of beta-lactam-induced delayed hypersensitivity and neutropenia during treatment of infective endocarditis. Arch Intern Med. 1999;159(6):607–615. doi:10.1001/archinte.159.6.607

26. Shah I, Kumar KS, Lerner AM. Agranulocytosis associated with chronic oral administration of cloxacillin for suppression of staphylococcal osteomyelitis. Am J Hematol. 1982;12(2):203–206. doi:10.1002/ajh.2830120213

27. Neftel KA, Hauser SP, Müller MR. Inhibition of granulopoiesis in vivo and in vitro by beta-lactam antibiotics. J Infect Dis. 1985;152(1):90–98. doi:10.1093/infdis/152.1.90

28. Weber D, Jenq RR, Peled JU, et al. Microbiota Disruption Induced by Early Use of Broad-Spectrum Antibiotics Is an Independent Risk Factor of Outcome after Allogeneic Stem Cell Transplantation. Biol Blood Marrow Transplant. 2017;23(5):845–852. doi:10.1016/j.bbmt.2017.02.006

29. Jenq RR, Taur Y, Devlin SM, et al. Intestinal Blautia Is Associated with Reduced Death from Graft-versus-Host Disease. Biol Blood Marrow Transplant. 2015;21(8):1373–1383. doi:10.1016/j.bbmt.2015.04.016

30. Peled JU, Devlin SM, Staffas A, et al. Intestinal Microbiota and Relapse After Hematopoietic-Cell Transplantation. J Clin Oncol. 2017;35(15):1650–1659. doi:10.1200/JCO.2016.70.3348

31. Peled JU, Gomes ALC, Devlin SM, et al. Microbiota as Predictor of Mortality in Allogeneic Hematopoietic-Cell Transplantation. N Engl J Med. 2020;382(9):822–834. doi:10.1056/NEJMoa1900623

32. Li J, Malouf C, Miles LA, Willis MB, Pietras EM, King KY. Chronic inflammation can transform the fate of normal and mutant hematopoietic stem cells. Exp Hematol. 2023;127:8–13. doi:10.1016/j.exphem.2023.08.008

33. Battini V, Mari A, Gringeri M, et al. Antibiotic-Induced Neutropenia in Pediatric Patients: New Insights From Pharmacoepidemiological Analyses and a Systematic Review. Front Pharmacol. 2022;13:877932. doi:10.3389/fphar.2022.877932

34. Solis K, Dehority W. Antibiotic-Induced Neutropenia During Treatment of Hematogenous Osteoarticular Infections in Otherwise Healthy Children. J Pediatr Pharmacol Ther. 2019;24(5):431–437. doi:10.5863/1551-6776-24.5.431

35. Zhang L, Huang Y, Zhou Y, Buckley T, Wang HH. Antibiotic Administration Routes Significantly Influence the Levels of Antibiotic Resistance in Gut Microbiota. Antimicrob Agents Chemother. 2013;57(8):3659–3666. doi:10.1128/AAC.00670-13

36. Ainonen S, Tejesvi MV, Mahmud MdR, et al. Antibiotics at birth and later antibiotic courses: effects on gut microbiota. Pediatr Res. 2022;91(1):154–162. doi:10.1038/s41390-021-01494-7

37. Xue L, Ding Y, Qin Q, et al. Assessment of the impact of intravenous antibiotics treatment on gut microbiota in patients: Clinical data from pre-and post-cardiac surgery. Front Cell Infect Microbiol. 2022;12:1043971. doi:10.3389/fcimb.2022.1043971

38. Babay HA, Twum-Danso K, Kambal AM, Al-Otaibi FE. Bloodstream infections in pediatric patients. Saudi Med J. 2005;26(10):1555–1561.

39. Kaplan SL. Implications of methicillin-resistant Staphylococcus aureus as a community-acquired pathogen in pediatric patients. Infect Dis Clin North Am. 2005;19(3):747–757. doi:10.1016/j.idc.2005.05.011

40. Vacca M, Celano G, Calabrese FM, Portincasa P, Gobbetti M, De Angelis M. The Controversial Role of Human Gut Lachnospiraceae. Microorganisms. 2020;8(4). doi:10.3390/microorganisms8040573

41. Hanachi M, Manichanh C, Schoenenberger A, et al. Altered host-gut microbes symbiosis in severely malnourished anorexia nervosa (AN) patients undergoing enteral nutrition: An explicative factor of functional intestinal disorders? Clin Nutr. 2019;38(5):2304–2310. doi:10.1016/j.clnu.2018.10.004

42. Ouaknine Krief J, Helly de Tauriers P, Dumenil C, et al. Role of antibiotic use, plasma citrulline and blood microbiome in advanced non-small cell lung cancer patients treated with nivolumab. J Immunother Cancer. 2019;7(1):176. doi:10.1186/s40425-019-0658-1

43. Niu K, Bai P, Zhang J, Feng X, Qiu F. Cytidine Alleviates Dyslipidemia and Modulates the Gut Microbiota Composition in ob/ob Mice. Nutrients. 2023;15(5). doi:10.3390/nu15051147

44. Singh V, Lee G, Son H, et al. Butyrate producers, “The Sentinel of Gut”: Their intestinal significance with and beyond butyrate, and prospective use as microbial therapeutics. Front Microbiol. 2022;13:1103836. doi:10.3389/fmicb.2022.1103836

45. Wan F, Deng FL, Chen L, et al. Long-term chemically protected sodium butyrate supplementation in broilers as an antibiotic alternative to dynamically modulate gut microbiota. Poult Sci. 2022;101(12):102221. doi:10.1016/j.psj.2022.102221

46. Hayase E, Hayase T, Jamal MA, et al. Mucus-degrading Bacteroides link carbapenems to aggravated graft-versus-host disease. Cell. 2022;185(20):3705–3719.e14. doi:10.1016/j.cell.2022.09.007

