## Supplemental Figures and Methods for "Antibiotic-associated neutropenia is marked by depletion of intestinal *Lachnospiraceae* in pediatric patients"

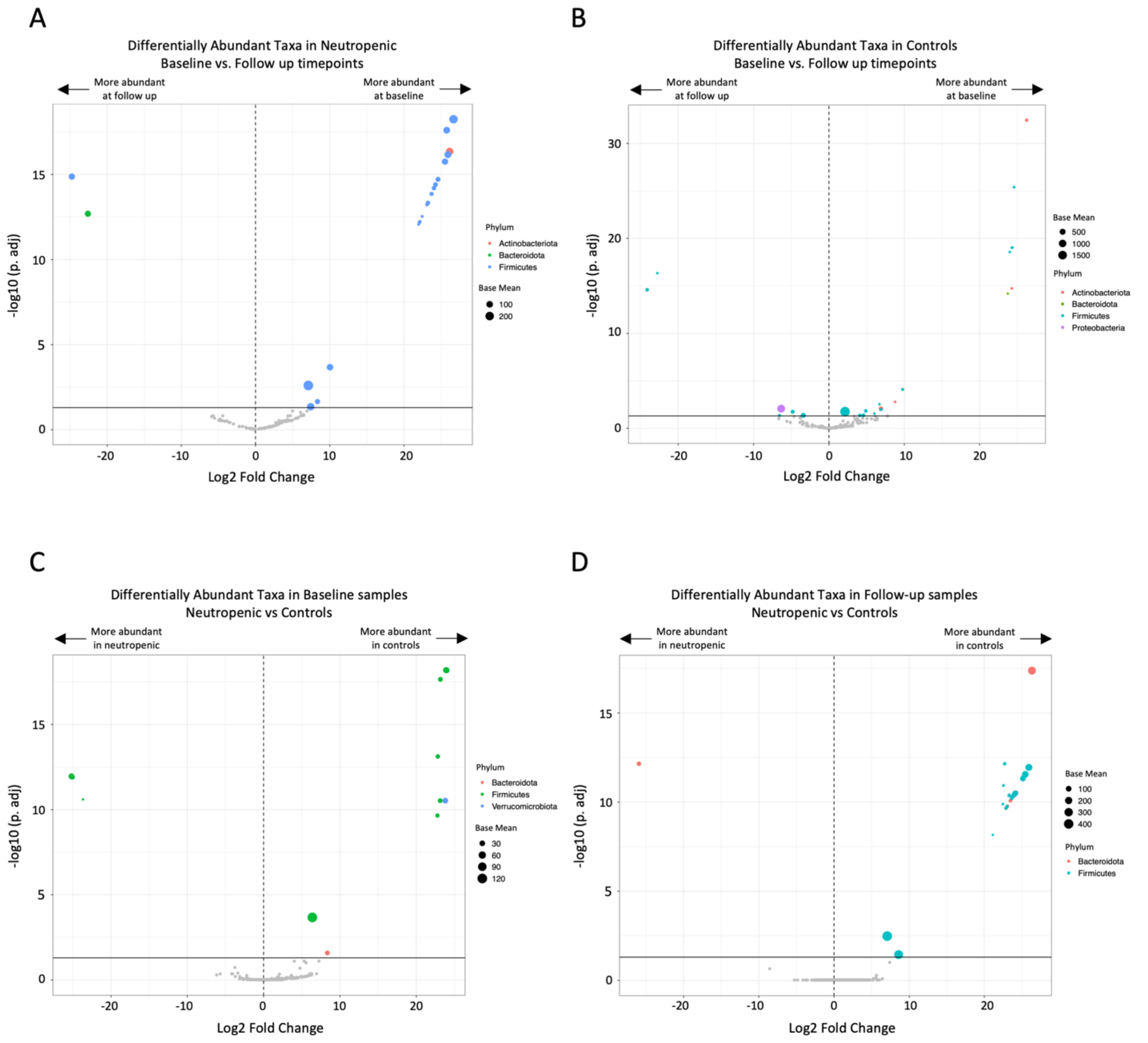


**Supplemental Figure 1. Several taxa were differentially abundant between groups and timepoints.** 16S rRNA analysis via DESeq2 was performed, volcano plots are shown and illustrate the four comparisons performed. Horizontal solid line indicates p.adj 0.05 of -log10 determined by Wald test. **A.** Differentially abundant taxa before and after antibiotic treatment in neutropenic subjects. **B.** Differentially abundant taxa before and after antibiotic treatment in control subjects. **C.** Differentially abundant taxa between neutropenic and control subjects before antibiotic treatment (baseline). **D.** Differentially abundant taxa between neutropenic and control subjects after antibiotic treatment (follow up). (n=10 neutropenic, 28 controls).

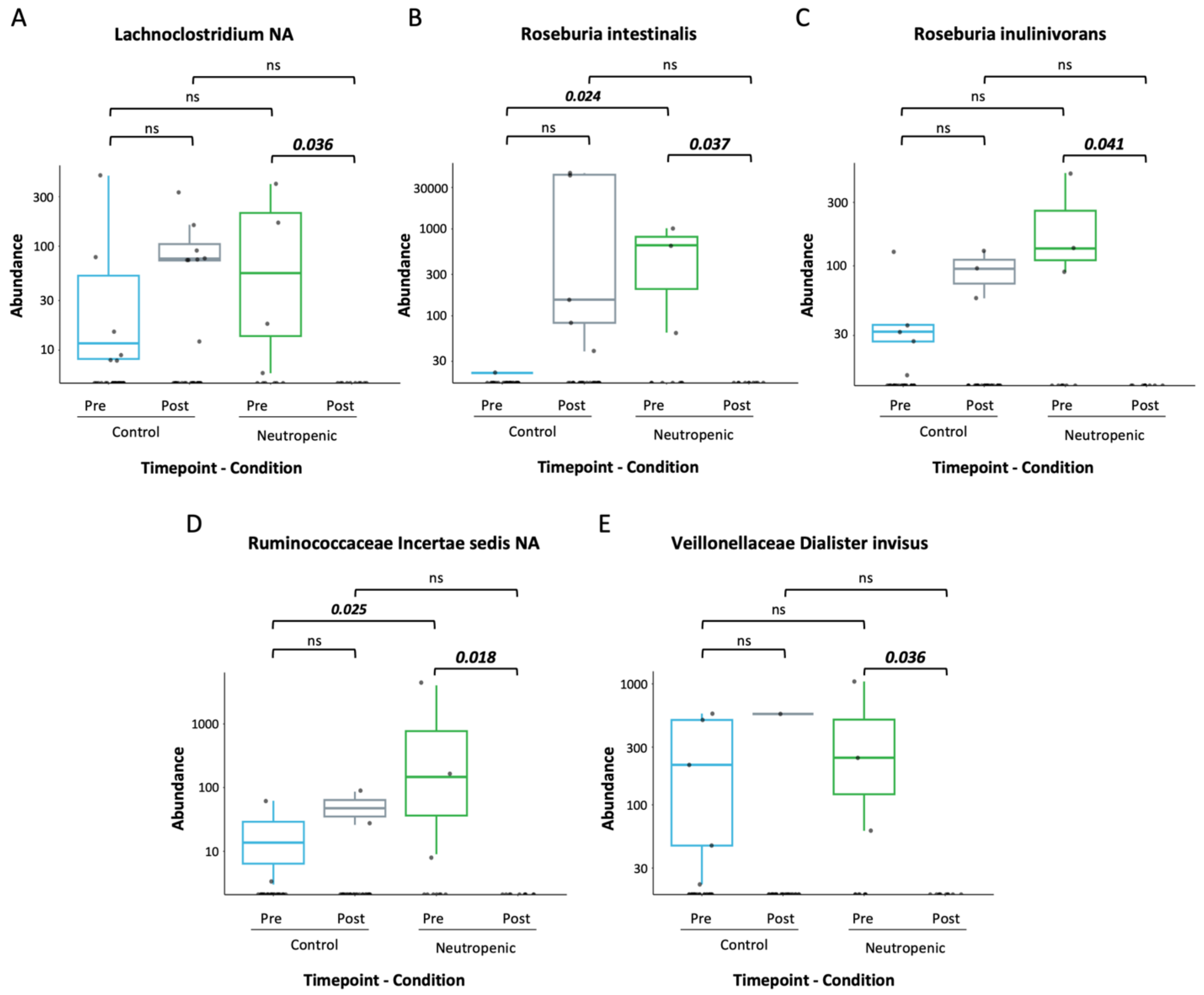


**Supplemental Figure 2. Several species demonstrated a trend towards lower abundance after antibiotic treatment only in neutropenic subjects.** Mean abundances by groups before and after antibiotic treatment. Statistical significance is defined by unadjusted $p$ <0.05 as determined by Dunn's multiple comparisons test (n=10 neutropenic, 28 controls).

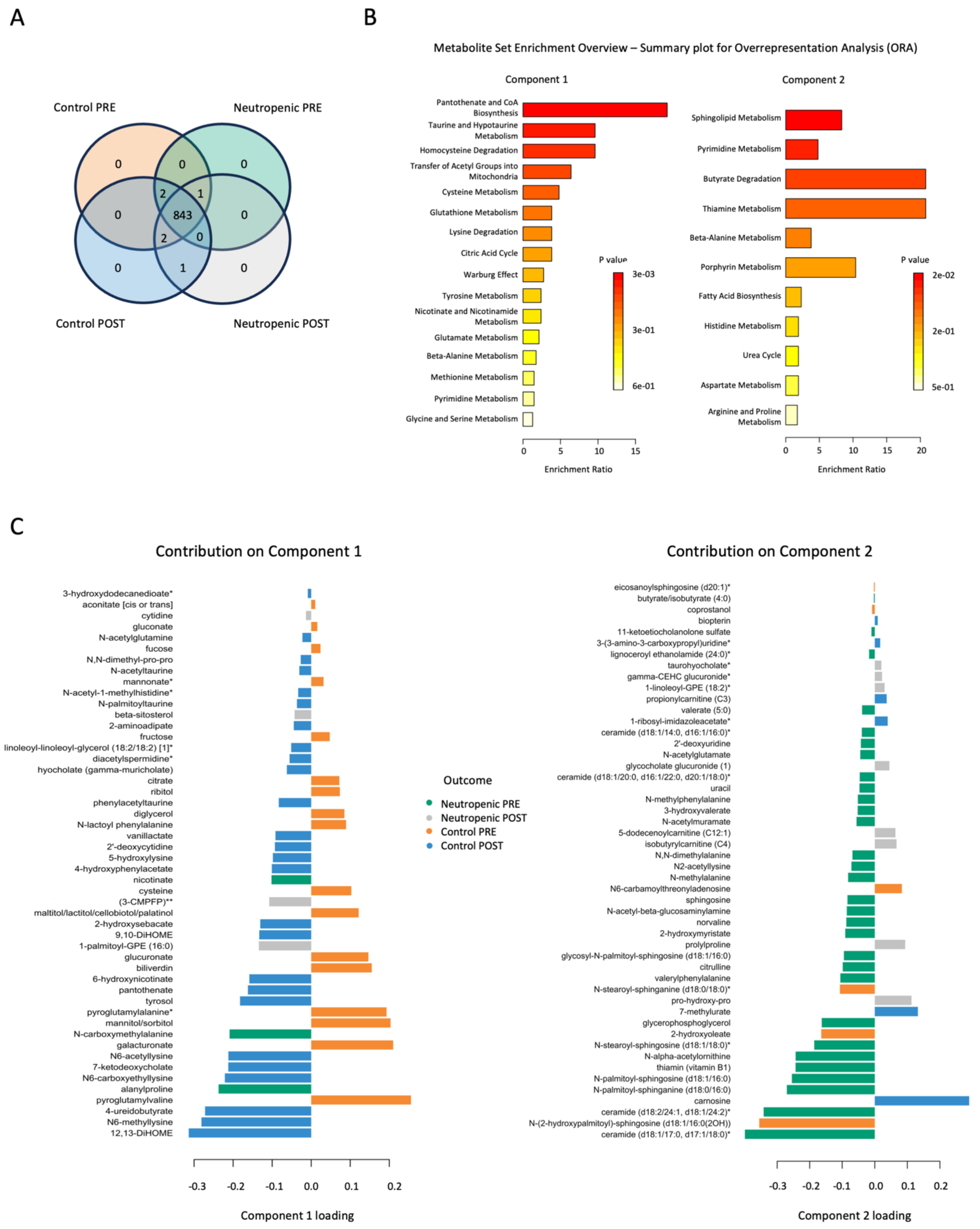


**Supplemental Figure 3. A broad range of metabolites were detectable in both neutropenic and control groups at both timepoints. A.** Most metabolites detected are present in samples from both groups before and after antibiotic treatment: Venn diagram shows 843 metabolites out of the 849 metabolites detected in all samples are present both before and after antibiotic treatment in subjects with and without neutropenia. **B.** Over-representation analysis (ORA) showed enrichment of metabolites involved in pantothenate and coenzyme A biosynthesis for the first component, whereas the second component demonstrated enrichment for metabolites involved in sphingolipid metabolism, pyrimidine metabolism, butyrate metabolism, and thiamine metabolism. **C.** Component loadings of metabolites contributing to sPLS-DA component 1 and 2 are shown. (n=10 neutropenic, 29 controls).

**Supplemental Methods**

**16s rRNA sequencing**

Total genomic DNA was extracted from human stool samples using the DNease PowerSoil Pro Kit (Qiagen) according to manufacturer's protocol. The 16S rDNA region was amplified by PCR and sequenced on the Illumina MiSeq platform (Table 1). Primers used for amplification contain adapters for MiSeq sequencing and single-index barcodes incorporated into the reverse primer so that resulting PCR products may be pooled and sequenced directly, targeting an average per/sample yield of 25,000 read pairs.

**Table 1**

| V REGION | FORWARD PRIMER | REVERSE PRIMER | PROTOCOL |
|---|---|---|---|
| v4 | 515F: GTGCCAGCMGCCGCGGTAA | 806R: GGACTACHVGGGTWTCTAAT | 2 x 250 bp |

Raw data files in binary base call (BCL) format were converted into FASTQs and demultiplexed based on the single-index barcodes using the Illumina 'bcl2fastq' software. Demultiplexed read pairs underwent an initial quality filtering using bbduk.sh (BBMap, version 38.82), removing Illumina adapters, PhiX reads and reads with a Phred quality score <15 and length <100 bp after trimming. Quality controlled reads were merged using bbmerge.sh4 with merge parameters: maxstrict=t, qtrim=t, trimq=15. Merged reads were further filtered via vsearch5 using parameters optimized for the appropriate 16S amplicon type (Table 2). Sequences were joined, trimmed to 150-bp reads, and denoised using Deblur through the QIIME2 pipeline using version 2022.2[1].

**Table 2**

| V REGION | MAX EXPECTED ERROR RATE | MIN LENGTH | MAX LENGTH |
|---|---|---|---|
| v4 | 0.05 | 252 bp | 254 bp |

**16S rRNA Sequencing Statistical analysis**

Sequencing Data used in this study are available at NCBI ENA under accession number PRJEB72348. Scripts are accessible at GitHub under project https://github.com/RachelRodgers/KKing-TINMAN-Collaboration

DADA2 was run in R version 4.0.0. The analysis of bacterial 16s RNA sequences was performed within R environment, version 4.0.5[2].

Raw amplicon analysis

The R package dada2 version 1.18.0 was used to obtain the table of exact amplicon sequence variants (ASVs) from raw amplicon sequence data[2]. ASVs were annotated using the SILVA v138.1 training set (https://benjjneb.github.io/dada2/training.html).

Differential taxa abundant analysis

The R package *DESeq2* version 1.18.0 was used to perform the taxa differential analysis between the different conditions[3].

Differential analysis of taxa counts abundance was assessed for the following groups and conditions:

- Neutropenic vs Control in the full cohort

- Neutropenic vs Control in samples post treatment only
- Neutropenic vs Control in samples pre-treatment only
- Post vs Pre-treatment samples in the Neutropenic population
- Post vs Pre-treatment samples in the Control population

The Deseq() function was run on the ASV table returned by the amplicon analysis, previously described, with the following parameters: *test = 'Wald', fitType='local'*

Taxa aggregation at the family level was calculated as the sum of all the read counts of the individual relative species. Dunn test was performed to compare mean abundance values for taxa of interest included in Table 3, Figure 4 and Supplemental Figure 2; as well as to compare the combined mean abundance values from the ten Lachnospiraceae species included in Table 3.

Alpha diversity analysis

Richness, Pielou's Evenness and Shannon Diversity were calculated with the microbiome package version 1.12.0.
The Mann-Whitney/Wilcoxon test was then used to compare statistical significance between the desired groups. The test was performed with the *wilcox.test()* function and $p$ values < 0.05 were considered to be statistically significant.

**Untargeted metabolomics**

**Sample Accessioning:** Following receipt, samples were inventoried and immediately stored at -80°C. Each sample received was accessioned into the Metabolon Laboratory Information Management System (LIMS) and was assigned by the LIMS a unique identifier that was associated with the original source identifier only. This identifier was used to track all sample handling, tasks, and results. The samples (and all derived aliquots) were tracked by the LIMS system. All portions of any sample were automatically assigned their own unique identifiers by the LIMS when a new task was created; the relationship of these samples was also tracked. All samples were maintained at -80°C until processed.

**Sample Preparation:** Samples were prepared using the automated MicroLab STAR® system (Hamilton). Several recovery standards were added prior to the first step in the extraction process for QC purposes. To remove protein, dissociate small molecules bound to protein or trapped in the precipitated protein matrix, and to recover chemically diverse metabolites, proteins were precipitated with methanol under vigorous shaking for 2 min (Glen Mills GenoGrinder 2000) followed by centrifugation. The resulting extract was divided into five fractions: two for analysis by two separate reverse phase (RP/UPLC-MS/MS) methods with positive ion mode electrospray ionization (ESI), one for analysis by RP/UPLC-MS/MS with negative ion mode ESI, one for analysis by HILIC/UPLC-MS/MS with negative ion mode ESI, and one sample was reserved for backup. Samples were placed briefly on a TurboVap® (Zymark) to remove the organic solvent. The sample extracts were stored overnight under nitrogen before preparation for analysis.

**QA/QC:** Several types of controls were analyzed in concert with the experimental samples: a pooled matrix sample generated by taking a small volume of each experimental sample (or alternatively, use of a pool of well-characterized human plasma) served as a technical replicate throughout the data set; extracted water samples served as process blanks; and a cocktail of QC standards that were carefully chosen not to interfere with the measurement of endogenous compounds were spiked into every analyzed sample, allowed instrument performance monitoring and aided chromatographic alignment. Tables 3 and 4 describe these QC samples and standards. Instrument variability was determined by calculating the median relative standard deviation (RSD) for the standards that were added to each sample prior to injection into the mass spectrometers.

Overall process variability was determined by calculating the median RSD for all endogenous metabolites (i.e., non-instrument standards) present in 100% of the pooled matrix samples. Experimental samples were randomized across the platform run with QC samples spaced evenly among the injections, as outlined in Figure 1.

Table 3: Description of Metabolon QC Samples

| Type | Description | Purpose |
|---|---|---|
| MTRX | Large pool of human plasma maintained by Metabolon that has been characterized extensively. | Assure that all aspects of the Metabolon process are operating within specifications. |
| CMTRX | Pool created by taking a small aliquot from every customer sample. | Assess the effect of a non-plasma matrix on the Metabolon process and distinguish biological variability from process variability. |
| PRCS | Aliquot of ultra-pure water | Process Blank used to assess the contribution to compound signals from the process. |
| SOLV | Aliquot of solvents used in extraction. | Solvent Blank used to segregate contamination sources in the extraction. |

Table 4: Metabolon QC Standards

| Type | Description | Purpose |
|---|---|---|
| RS | Recovery Standard | Assess variability and verify performance of extraction and instrumentation. |
| IS | Internal Standard | Assess variability and performance of instrument. |

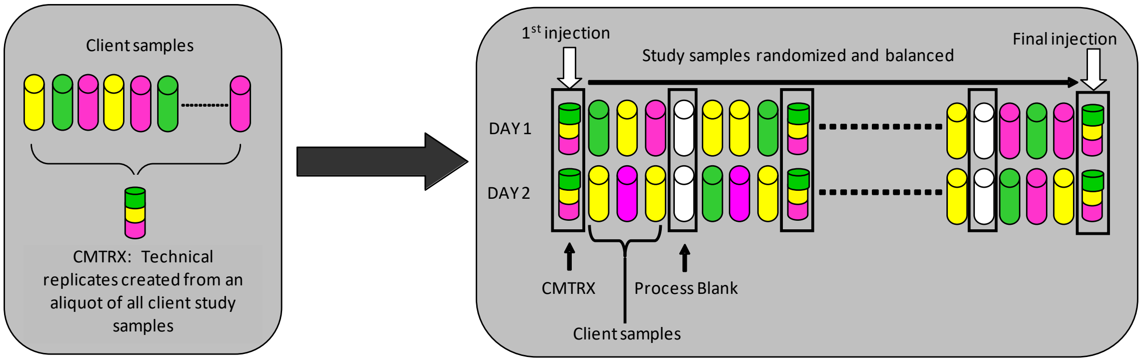


Figure 1. Preparation of technical replicates. A small aliquot of each sample (colored cylinders) is pooled to create a CMTRX technical replicate sample (multi-colored cylinder), which is then injected periodically throughout the platform run. Variability among consistently detected biochemicals can be used to calculate an estimate of overall process and platform variability.

**Ultrahigh Performance Liquid Chromatography-Tandem Mass Spectroscopy (UPLC-MS/MS):** All methods utilized a Waters ACQUITY ultra-performance liquid chromatography (UPLC) and a Thermo Scientific Q-Exactive high resolution/accurate mass spectrometer interfaced with a heated electrospray ionization (HESI-II) source and Orbitrap mass analyzer operated at 35,000 mass resolution. The sample extract was dried then reconstituted in solvents compatible to each of the four methods. Each reconstitution solvent contained a series of standards at fixed concentrations to ensure injection and chromatographic consistency. One aliquot was analyzed using acidic positive ion conditions, chromatographically optimized for more hydrophilic compounds. In this method, the extract was gradient eluted from a C18 column

(Waters UPLC BEH C18-2.1x100 mm, 1.7 µm) using water and methanol, containing 0.05% perfluoropentanoic acid (PFPA) and 0.1% formic acid (FA). Another aliquot was also analyzed using acidic positive ion conditions; however, it was chromatographically optimized for more hydrophobic compounds. In this method, the extract was gradient eluted from the same afore mentioned C18 column using methanol, acetonitrile, water, 0.05% PFPA and 0.01% FA and was operated at an overall higher organic content. Another aliquot was analyzed using basic negative ion optimized conditions using a separate dedicated C18 column. The basic extracts were gradient eluted from the column using methanol and water, however with 6.5mM Ammonium Bicarbonate at pH 8. The fourth aliquot was analyzed via negative ionization following elution from a HILIC column (Waters UPLC BEH Amide 2.1x150 mm, 1.7 µm) using a gradient consisting of water and acetonitrile with 10mM Ammonium Formate, pH 10.8. The MS analysis alternated between MS and data-dependent MSn scans using dynamic exclusion. The scan range varied slighted between methods but covered 70-1000 m/z. Raw data files are archived and extracted as described below.

**Bioinformatics:** The informatics system consisted of four major components, the Laboratory Information Management System (LIMS), the data extraction and peak-identification software, data processing tools for QC and compound identification, and a collection of information interpretation and visualization tools for use by data analysts. The hardware and software foundations for these informatics components were the LAN backbone, and a database server running Oracle 10.2.0.1 Enterprise Edition.

**LIMS:** The purpose of the Metabolon LIMS system was to enable fully auditable laboratory automation through a secure, easy to use, and highly specialized system. The scope of the Metabolon LIMS system encompasses sample accessioning, sample preparation and instrumental analysis and reporting and advanced data analysis. All of the subsequent software systems are grounded in the LIMS data structures. It has been modified to leverage and interface with the in-house information extraction and data visualization systems, as well as third party instrumentation and data analysis software.

**Data Extraction and Compound Identification:** Raw data was extracted, peak-identified and QC processed using Metabolon's hardware and software.

**Curation:** A variety of curation procedures were carried out by Metabolon to ensure accurate and consistent identification of true chemical entities, and to remove those representing system artifacts, mis-assignments, and background noise. Library matches for each compound were checked for each sample and corrected if necessary.

**Metabolite Quantification and Data Normalization:** Peaks were quantified using area-under-the-curve. In certain instances, biochemical data may have been normalized to an additional factor (e.g., cell counts, total protein as determined by Bradford assay, osmolality, etc.) to account for differences in metabolite levels due to differences in the amount of material present in each sample.

**Statistical Methods and Analysis**

Following imputation of missing values, data were auto scaled prior to analysis[4]. To determine how metabolite abundances changed over the course of therapy in neutropenic patients and controls, we calculated log2-scale mean fold change in each group as the mean post-treatment divided by the mean pre-treatment abundance and compared pre- and post-treatment metabolite abundances by Quade test[5], which accounts for subject effects. We visualized these results in a volcano plot. We then performed Partial Least Squares-Discriminant Analysis (PLS-DA) using the 'mixOmics' package v6.26.0 in R v4.2.1 to evaluate whether the metabolomes of neutropenic patients and controls differed at each time point. Metabolite set enrichment analysis (MSEA) was performed using MetaboAnalyst 5.0 web interface, comparing post-antibiotic therapy samples between neutropenic and control subjects. The Small Molecule Pathway Database (SMPBD) was used as reference databases.

To determine whether the metabolites that defined PLS-DA components one and two were associated with neutropenia, we computed logistic regression models where the outcome was the odds ratio (OR) and 95% confidence interval (CI) of neutropenia for a one-standard deviation increase in abundance. First, we estimated the OR and CI of neutropenia associated with a one standard deviation increase in the abundance of each metabolite at the pre-treatment timepoint. Our objective was to identify metabolites that may serve as biomarkers of neutropenia risk at baseline. To understand how changes in the metabolome across therapy were associated with neutropenia, we separately estimated the OR and CI of neutropenia for each one standard deviation increase in the abundance of the index metabolite from the pre-treatment to the post-treatment timepoint, adjusting for its baseline abundance. To characterize the biological processes encoded by the first two PLS-DA components, we performed over-representation analysis (ORA) in MetaboAnalyst 5.0. Metabolites that defined these components were compared to a background set of all measured endogenous or bacterial/fungal metabolites to determine whether there was evidence of enrichment for metabolites mapped to normal human metabolic pathways defined by SMPDB.

The R script used for metabolomics analysis is available at https://github.com/orgs/BCM-TCH-EpiCenter/repositories

**References**


1. Amir A, McDonald D, Navas-Molina JA, et al. Deblur Rapidly Resolves Single-Nucleotide Community Sequence Patterns. mSystems. 2017;2(2).
2. Team RC. R: A language and environment for statistical computing. R Foundation for Statistical Computing. Vienna, Austria; 2021.
3. Callahan BJ, Sankaran K, Fukuyama JA, McMurdie PJ, Holmes SP. Bioconductor Workflow for Microbiome Data Analysis: from raw reads to community analyses. F1000Res. 2016;5:1492.
4. Van den Berg RA, Hoefsloot HC, Westerhuis JA, Smilde AK, van der Werf MJ. Centering, scaling, and transformations: improving the biological information content of metabolomics data. BMC Genomics. 2006;7:142.
5. Campbell RA. A comparison of the Quade and Friedman tests to the unbalanced two-way analysis of variance with biomedical data. Comput Biol Med. 1988;18(6):441-7.
